# Large-Scale Identification of Novel Protein Biomarkers and Therapeutic Targets in Heart and Brain Disease

**DOI:** 10.64898/2026.01.26.26344874

**Authors:** Chao Wu, Dantong Li, Sumeet A. Khetarpal, Zixun Yuan, Shengyuan Huang, Justin Ralph Baldovino Guerra, Chunyu Li, Qiulian Zhou, Meixi Quan, Jiaqi He, Meng Wang, Huiying Liang, Anthony Rosenzweig

**Author notes:** These authors contribute equally to this manuscript (Chao Wu, Dantong Li).

## Abstract

Neurological complications frequently impact morbidity, mortality, and quality of life in patients with cardiovascular disease, yet the biological mediators connecting cardiovascular and neurological disease are poorly understood. Leveraging data from 53,014 individuals with plasma proteomic profiles and 50,228 with cardiac and brain MRI from the UK Biobank, we systematically identified circulating proteins correlated with MRI imaging-derived phenotypes (IDPs) (404 proteins with cardiac IDPs; 76 with brain IDPs; 37 with both). Identified proteins were remarkably enriched for biomarkers and mediators of disease in one or both organs. Expression analyses suggested these proteins largely originate from fibroblasts, smooth muscle cells, and macrophages in the arterial vasculature. Pathway analyses highlighted cytokine and vasculature-related processes for cardiac IDPs-associated proteins and extracellular matrix pathways in brain IDPs-associated proteins. Mendelian Randomization and genetic co-localization supported causal roles for most (>63%) of the proteins in disease pathogenesis in one or both organs. Over 90% of the implicated candidates have not previously been established as clinical biomarkers or therapeutic targets. These studies underscore the value of large-scale integrated multi-organ datasets, including plasma proteomics, imaging-derived endophenotypes, and genetics, in unraveling complex disease pathobiology, highlight the close connections between heart and brain disease, and provide a catalog of hundreds of novel candidate biomarkers and therapeutic targets.

## INTRODUCTION

Growing evidence suggests the heart and brain are intimately connected, with important implications for human health. Neurological complications occur frequently in patients with cardiovascular disease and are key determinants of clinical outcomes and quality of life. Thus, understanding this connection is particularly important for patients with cardiovascular disease.

Children with congenital heart disease have a higher risk of neurodevelopmental abnormalities, at least in part due to the prevalence of mutations in genes common to heart and brain development^1^. Moreover, there is a strong association between heart failure (HF)^2^ and neurodegenerative disease^3,4^, and the risk factors^5,6^, predisposing genetic factors^7–9^, and mitigating factors^10,11^ significantly overlap.

Moreover, each organ profoundly impacts the other. Most obvious are the adverse neurological sequelae caused by alterations in cardiac rhythm or function. However, more subtle crosstalk between the organs is increasingly recognized. In murine models, optogenetically-induced tachycardia amplified anxiety manifestations via the posterior insular cortex^12^, resonating with subjective sensations reported by arrhythmia patients. Structural artery-brain circuits have been shown to modulate disease progression and plaque stability in murine models of atherosclerosis^13^, reflecting interactions among the nervous, immune, and vascular systems termed neuroimmune cardiovascular interfaces. In humans, cardiac complications of acute stroke occur even absent prior cardiovascular disease^14^. This neuro-cardiac signaling^15^ is recapitulated in murine models where cardiac injury after stroke appears mediated by activated myeloid immune cells^16^. Even more dramatically, severe cardiomyopathy can be induced by emotional stress^17^ in association with alterations in brain functional magnetic resonance imaging (fMRI)^18^. Thus, bidirectional heart-brain signaling has important health consequences and its molecular mechanisms are not well understood.

Zhao and colleagues recently used MRIs from the UK Biobank (UKB) to identify associations between cardiac and brain imaging traits^19^. Genetic analyses supported bidirectional connections and potentially causal relationships between heart and brain traits^19^. However, the mediators and mechanisms connecting the heart and brain, as well as the broader implications for disease, remained unclear. Here, we used targeted plasma proteomic data and MRI imaging data in 53,014 and 50,228 UKB participants, respectively, to investigate the role of circulating proteins, inherently accessible as potential biomarkers or therapeutic targets, in heart-brain connections and disease. Many plasma proteins associated with MRI imaging derived phenotypes (IDPs) in the heart, brain, or both. Strikingly, most IDP-associated proteins were predictive of incident heart and/or brain disease and genetic analyses employing Mendelian randomization (MR) and genetic colocalization suggested many have potentially causal roles. Many proteins associated with IDPs in one organ had previously unappreciated associations with disease in that organ and the other. Bioinformatic analyses revealed inflammatory and extracellular matrix as dominant pathways, and implicated arterial cells as the likely source and responder for many proteins. Taken together, these data underscore the key role of circulating proteins in connecting heart and brain biology and disease, and provide an atlas of genetically validated mediators, clinical biomarkers, and potential therapeutic targets.

## RESULTS

### Participants

Data from 53,014 UKB participants with Olink Explore 3072 plasma proteomics data from the Pharma Proteomics Project (UKB-PPP)^20^ and 50,228 participants with cardiac (CMR) and brain (BMR) MRI scans were analyzed (Fig. 1a). IDPs were obtained from 47,453 BMR and 39,676 CMR scans. The mean age of participants in the proteomics cohort was 57.3 ± 8.2 years, while the MRI cohort had a mean age of 64.9 ± 7.8 years. Females comprised 53.9% of the proteomics cohort and 52.1% of the MRI cohort, and the majority of participants were White British (93.6% and 96.6%, respectively). Additional baseline characteristics of the study population are provided in Extended Data Table 1.

**Fig. 1.**
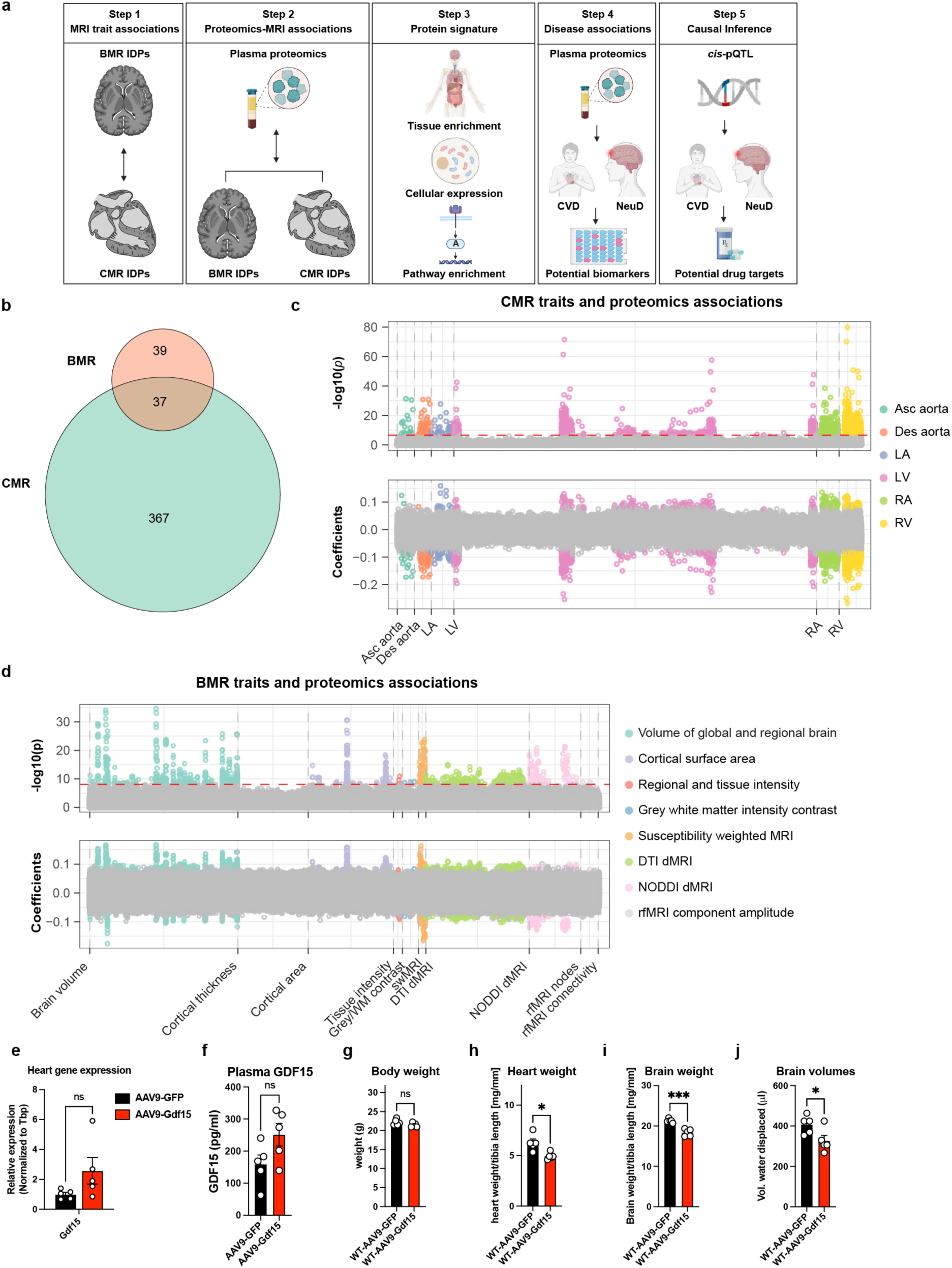
Correlations between proteomics and MRI traits. **a.** Overall study workflow, created by BioRender. **b**. Number of proteins significantly associated with cardiac MRI (CMR) traits, brain MRI (BMR) traits, or both (Bonferroni-corrected). **c-d**. Manhattan plot illustrating correlations between plasma proteins and CMR traits (**c**) or BMR traits (**d**). The y-axis shows −log_10_(*p*) values, and correlation coefficients are indicated for significant associations. The red line denotes the Bonferroni-corrected threshold (*p*<2.09×10^−7^ for CMR traits and *p*<7.70×10^−9^ for BMR traits); gray points represent non-significant correlations. **e**, Cardiac *Gdf15* expression in mice injected with AAV9-cTnT-*Gdf15* or AAV9-cTnT-GFP control. **f**, Plasma GDF15 concentrations in the same mice (n = 5 per group). Data are presented as mean ± SEM, ns, not significant. **g-i**, Weights of whole body (**g**), heart (**h**), and brain (**i**). **j**, Brain volume measured by water displacement. n = 5 per group. Data are presented as mean ± SEM; ns, not significant; \**p* < 0.05, \*\*\**p* < 0.001. cis-pQTL, cis-protein quantitative trait locus; WM, white matter; swMRI, susceptibility-weighted MRI; DTI, diffusion tensor imaging; NODDI, neurite orientation dispersion and density imaging; dMRI, diffusion-weighted MRI; rfMRI, resting-state functional MRI; LV, left ventricle; LA, left atrium; RV, right ventricle; RA, right atrium; Asc aorta, ascending aorta; Des aorta, descending aorta; CVD, cardiovascular diseases; NeuD, neurological diseases; IDP, imaging derived phenotype.

### Structural and functional heart-brain interactions

Pairwise univariate analyses of 82 CMR and 2,222 BMR IDPs in 32,859 participants, excluding task fMRI traits due to lower reproducibility^21^, confirmed a strong correlation between heart-brain IDPs^19^ with an expanded set of BMR IDPs. After adjusting for confounders (see Methods), we identified 15,793 significant associations after Bonferroni correction (Extended Data Fig. 1a).

Many associations (38.1%) involved left ventricular (LV) phenotypes and diffusion tensor imaging (DTI) metrics of white matter integrity, while 13.6% involved neurite orientation dispersion and density imaging (NODDI) measures, which reflect microstructural features. For example, LV myocardial mass correlated with mean diffusivity in the left external capsule (*r*=0.14, *p*=5.25×10^−140^). Right ventricular (RV) measures were linked to global and regional brain volumes in 223 associations (1.4%). Ascending and descending aortic distensibility both showed strong negative correlations with white matter hyperintensity (WMH) volume, a potential indicator of neuropathology, with *r* ≈ −0.10 to −0.11, *p*<1×10^−62^, whereas aortic area measures were positively correlated with WMH (Extended Data Fig. 1b,c).

Our findings align well with the prior report^19,22^, including positive correlations between global myocardial wall thickness and subcortical volumes (e.g., putamen) and negative correlations between LV mass and fractional anisotropy. Notably, we additionally observed that LV myocardial mass was positively correlated with hippocampal volume but inversely associated with hippocampal microstructure, as reflected by intracellular volume fraction. This divergence between macrostructural and microstructural hippocampal measures suggests that HF-related cardiac remodeling may differentially impact hippocampal integrity. Importantly, these CMR-BMR IDP associations establish a necessary analytic framework for the subsequent multi-omic, inter-organ investigations. By reaffirming and extending known heart-brain imaging relationships using updated IDPs, this analysis provides a robust foundation for interrogating the contribution of circulating proteins to heart-brain interactions.

To further assess the cognitive significance of these brain MRI IDPs, we conducted univariable correlation analyses between 2,222 brain IDPs and eight cognitive assessments. These assessments were categorized into domains of numeric memory, prospective memory, and fluid intelligence. The analyses revealed strong associations between multiple BMR IDPs and cognitive performance. Out of the 17,766 correlations tested, 1,107 pairs met the Bonferroni threshold (*p*<2.81×10^−6^). Notably, 538 BMR traits were found to be significantly correlated with three different cognition categories. The complete results are presented in Extended Data Fig. 2, and support the potential functional significance of even subclinical brain MRI imaging traits.

### Correlations between proteomics and MRI phenotypes

Using data from 6,412 participants who had both proteomic measurements (2,922 Olink assays) and CMR/BMR data, we examined pairwise correlations between plasma proteins and IDPs.

2,097 significant correlations (*p*<2.10×10^−7^, Bonferroni-corrected) linked 404 proteins to 42 cardiac traits (Fig. 1b,c). Right and left ventricular phenotypes each correlated with nearly 300 of these proteins, including notable associations with fatty acid-binding protein-4 and leptin across all four chambers and both ascending/descending aorta. Renin, the apex of the renin-angiotensin-aldosterone system (RAAS), is associated with 25 CMR traits, consistent with its established roles in fluid homeostasis and vascular tone.

Eight hundred twenty-seven significant associations (*p*<7.70×10^−9^) emerged between 76 proteins and 357 brain traits (Fig. 1b,d). Prominent examples included oligodendrocyte-myelin glycoprotein, brevican core protein (BCAN), and neurocan core protein (NCAN), which influence central nervous system (CNS) architecture. Glial fibrillary acidic protein, implicated in neuroinflammation^23,24^, showed the most BMR correlations (139). Overall size and volumetric measures (e.g., brain segmentation volume to intracranial volume, total intracranial volume) had the largest number of protein associations, hinting at potential roles in neurodegenerative or developmental pathways.

37 proteins correlated significantly with both CMR and BMR traits, suggesting shared biological mechanisms

Although our primary objective was to comprehensively characterize associations between the circulating proteome and MRI-derived phenotypes, we recognize that these relationships may be influenced by multiple covariates. To evaluate the robustness of our findings, we performed two additional sensitivity analyses with progressively more extensive covariate adjustment. Importantly, because some covariates may lie on the causal pathway linking proteomic variation to imaging phenotypes, the primary model was intentionally specified to minimize overadjustment bias, with sensitivity analyses used to confirm robustness.

In the primary discovery model, we adjusted for technical factors and basic demographic variables to maximize sensitivity for detecting broad proteome-MRI associations. We then applied a sequential adjustment strategy to distinguish baseline associations from those independent of socioeconomic and cardiometabolic factors. First, we additionally adjusted for two socioeconomic variables: Townsend deprivation index and educational attainment. Under this model, over 96% (389/404) of protein-CMR trait associations identified in the primary analysis remained statistically significant, as did over 99% (75/76) of protein-BMR trait associations.

We next implemented a more stringently adjusted model that further incorporated cardiometabolic and lifestyle covariates, including systolic blood pressure, dyslipidemia, type 2 diabetes, smoking status, and alcohol consumption. Despite this extensive adjustment, substantial overlap with the primary results persisted and the overall conclusions were unchanged. Specifically, 76% (306/404) of protein-CMR trait associations and 63% (48/76) of protein-BMR trait associations remained significant. Collectively, these results demonstrate that the observed proteome-MRI associations are robust and not primarily driven by socioeconomic status, cardiometabolic risk, or lifestyle factors.

### Plasma proteins as mediators of heart-brain interactions

We used mediation analysis to assess whether circulating proteins provide pathways linking structural changes in one organ to changes in the other (Extended Data Fig. 3a,b). Among 37 proteins correlated with IDPs in both organs, 17 were associated with both paired traits, enabling mediation analysis of 21 cardiac and 72 brain traits. A protein was deemed a significant mediator if its indirect (protein-driven) and total effects were both significant at 5% false discovery rate (FDR), with consistent directionality.

Ten of these 17 proteins mediated significant relationships from 28 brain to 18 heart IDPs, and 9 of 17 mediated relationships from heart to brain IDPs. Notably, growth/differentiation factor 15 (GDF15) mediated 22 of 27 cross-organ associations. For example, GDF15, previously linked to ischemia^25^, cardiovascular outcomes^26^, and inflammation^27^, explained 29.4% of the link between mean isotropic volume fraction in the right fornix crescent/stria terminalis, involved in memory consolidation and stress regulation, and RV end-diastolic volume. GDF15 inhibitors being developed for other clinical indications could potentially interrupt this cascade^28^. Cadherin-related family member 2 (CDHR2) accounted for 30.8% of the link between right atrial (RA) volume and left putamen magnetic susceptibility, an indicator of subcortical iron or myelin changes seen with ischemic injury and inflammation. NCAN mediated 27.8% of the relationship between LV myocardial mass and mean perpendicular diffusivity in the genu of the corpus callosum, suggesting effects on axonal or myelin integrity. Collectively, these proteins (Extended Data Fig. 2c,d) highlight potential molecular mechanisms through which structural changes in each organ may mediate changes in the other.

Given that cross-sectional omics analyses are inherently susceptible to confounding and reverse causation, we conducted Mendelian randomization (MR) analyses to provide complementary genetic support for causal inference. Because large-scale GWAS summary statistics are not currently available for all cardiac and brain MRI traits, these analyses were necessarily focused on nine protein mediator candidates identified in the mediation framework and their corresponding significant cardiac-brain MRI trait pairs. Genome-wide significant *cis*-pQTLs were used as instrumental variables and were obtained from the UKB-PPP^20^, while GWAS summary statistics for brain MRI structural phenotypes were sourced from the MRC IEU OpenGWAS repository^29,30^.

We implemented a two-sample MR framework incorporating twelve complementary methods to account for horizontal pleiotropy and heterogeneity, and we additionally applied Steiger directionality testing to confirm the orientation of effects from protein exposure to imaging outcome. Several candidate proteins demonstrated genetic evidence consistent with potential causal effects on brain structural phenotypes. For example, NCAN showed inverse associations with thalamic volume (left thalamus*: β* = −0.15, *p* = 2.9 × 10⁻⁴; right thalamus: *β* = −0.14, *p* = 7.2 × 10⁻⁴). In addition, GDF15 was positively associated with a diffusion MRI-based Tract-Based Spatial Statistics measure reflecting radial diffusivity-related microstructural integrity in the left fornix crus and stria terminalis (*β* = 0.07, *p* = 4.3 × 10⁻³), consistent with increased microstructural vulnerability in limbic white matter.

While these MR results provide supportive genetic for a subset of protein mediators influencing brain structural variation, we emphasize that the primary aim of these analyses is to complement the mediation framework rather than to provide definitive causal proof. In the context of heart-brain interactions, the absence of statistically significant MR evidence for some protein mediators should therefore not be interpreted as evidence against their biological relevance. Such proteins may still represent meaningful mediators of cardiac-brain imaging associations, reflecting downstream, shared, or environmentally-modulated biological processes rather than direct genetically driven causal effects.

Given its consistent associations in our analyses, we next investigated the role of GDF15 in an animal model. Six- to eight-week-old wild-type male mice were injected with AAV9-cTnT-GFP or AAV9-cTnT-Gdf15 (5×10^11^ vector genomes per mouse). After 4 weeks of AAV expression, mice were subjected to voluntary wheel running for 7 days. Then, hearts and brains were harvested for analysis. Cardiac-specific overexpression of Gdf15 (Fig. 1e,f) led to reductions in heart and brain weights, as well as decreased brain volumes (Fig. 1g-j), compared with control animals. These findings parallel our human population findings, where higher GDF15 levels were associated with lower RV end-diastolic volume (*r* = −0.12, *p*=1.03×10^−16^), LV end-diastolic volume (*r*=-0.08, *p*=6.9 ×10^−9^), and total brain volume (*r*=-0.09, *p*=7.57×10^−11^), as well as other structural and functional brain measures. Together, these experimental and population-based results reinforce each other, supporting our mediation analysis inference that GDF15 may contribute to the link between reduced LV end-diastolic volume and smaller total brain volumes (mediation proportion=22.2%).

### Tissue enrichment and cellular expression of imaging trait-associated proteins

We investigated the likely sources of proteins correlated with IDPs using the Genotype-Tissue Expression (GTEx) framework^31,32^: proteins scoring ≥4 in one tissue and <2.5 in all others were considered “tissue-specific” and “tissue-enriched” if the top tissue scored ≥2.5 but was not tissue-specific. 401 of the 404 CMR-associated proteins had tissue enrichment data. Of these, 188 were tissue-specific or enriched, 58 were broadly expressed, and 155 lacked distinct tissue enrichment (Fig. 2a). 75 of 76 BMR-associated proteins had enrichment data: 44 were tissue-specific/enriched; 6 were broadly expressed; and 25 lacked distinct enrichment. These proportions are similar to the Olink panel overall, where ∼46% are tissue-specific/enriched (Extended Data Fig. 3a).

**Fig. 2.**
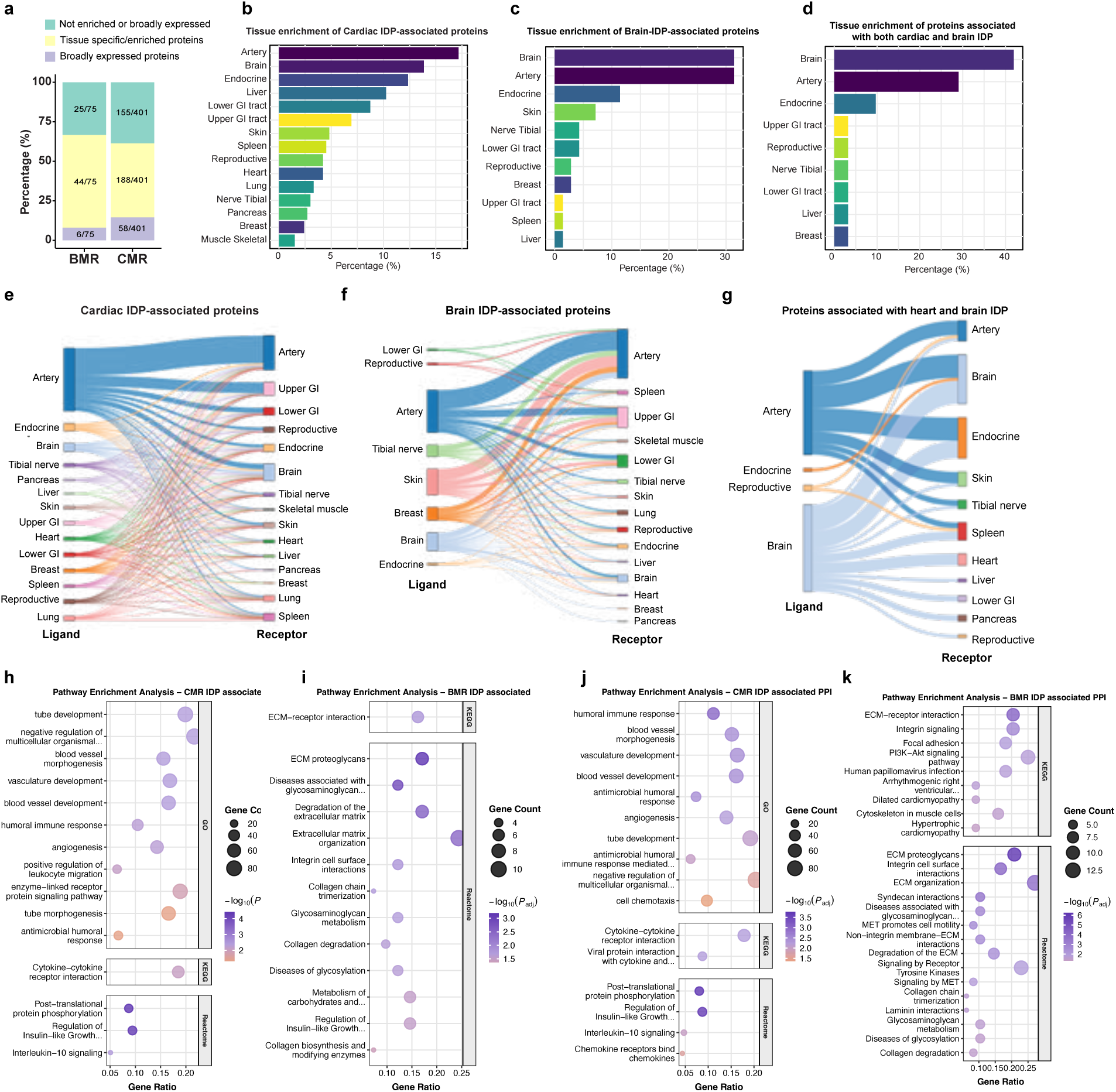
Tissue and pathway enrichment of MRI trait-associated proteins. **a.** Distribution of tissue enrichment (tissue-specific, tissue-enriched, broadly expressed, unknown) for cardiac IDP and brain IDP-associated proteins. **b–d**. Tissue enrichment across 15 consolidated tissue groups for proteins significantly associated with cardiac IDP (**b**), brain IDP (**c**), or both (**d**). **e–g**. Plasma protein tissue enrichment and corresponding receptor tissue enrichment for proteins associated with cardiac IDP (**e**), brain IDP (**f**), or both (**g**). **h–i.** Gene set enrichment analysis of pathways for CMR IDP-associated (**h**) and BMR IDP-associated (**i**) plasma proteins. **j–k,** Pathway enrichment after expanding each protein set with protein-protein interaction (PPI) neighbors for CMR (**j**) and BMR (**k**) IDP–associated proteins. GI, gastrointestinal; KEGG, Kyoto Encyclopedia of Genes and Genomes; PPI, protein-protein interactions.

We grouped the 32 GTEx tissues into 15 closely related categories. Notably, among tissue-specific/enriched proteins associated with CMR traits, the most common likely tissue sources were artery (∼17%), brain (∼14%), and endocrine (∼12%), whereas the heart contributed ∼4% (Fig. 2b, Extended Data Fig. 5a). Similarly, for BMR-associated proteins, artery (∼31%), brain (∼31%), and endocrine (∼11%) were most prevalent (Fig. 2c, Extended Data Fig. 5b). 23 of 37 proteins linked to both CMR and BMR traits were tissue-specific/enriched, largely in brain (∼41%) and artery (∼29%) (Fig. 2d, Extended Data Fig. 4d,e). The high prevalence of arterial proteins compares to ∼11% represented on the Olink platform used (Extended Data Fig. 4b,c), underscoring the importance of the vasculature in remodeling of both the heart and brain. While tissue enrichment analyses provide valuable biological context, they are inherently limited by reliance on reference datasets and cannot definitively establish tissue of origin or fully exclude residual effects related to protein abundance or detectability. Accordingly, the enrichment results presented here should be interpreted as contextual evidence of potential tissue relevance rather than definitive attribution, and will require targeted experimental and biological validation to confirm tissue sources and functional roles of key protein candidates.

To dissect cellular sources of artery-enriched proteins, we analyzed single-cell RNA-sequencing (scRNAseq) data from aorta (GSE15546829)^33^ and carotid (GSE25390330)^34^ (Extended Data Figs. 6, 7a,b). Of artery-enriched cardiac IDP-associated proteins represented in the aortic scRNAseq data, 72% were expressed in fibroblasts, 68% in smooth muscle cells (SMCs), 44% in monocytes/macrophages, 24% in endothelial cells (ECs). Among artery-enriched brain IDP-associated proteins identified in aortic scRNAseq, 6 of 7 were expressed in SMCs, 5/7 in fibroblasts, 5/7 in monocytes/macrophages, and 2 in ECs (Extended Data Fig. 7c,d). Comparable patterns emerged in carotid tissue (Extended Data Fig. 7e,f), underscoring fibroblasts, smooth muscle cells, and monocytes/macrophages as key cell types contributing artery-enriched proteins linked to heart and brain IDPs.

### Receptor distribution for imaging trait-associated proteins

We used OmniPath^35,36^ to identify the proteins’ receptors, grouping them by tissue enrichment (Fig. 2e-g). For proteins associated with CMR traits, by far the most frequent ligand-receptor tissue interactions were artery-to-artery (312), followed by brain-to-brain (34) and brain-to-heart (7) (Fig. 2e, Extended Data Fig. 5c). BMR trait-associated proteins showed a similar pattern (Fig. 2f, Extended Data Fig. 5d). Among proteins linked to both CMR and BMR traits, the brain was a prominent source and target of circulating ligands (Fig. 2g, Extended Data Fig. 4f). These patterns suggest that arteries serve as both a major source and target of proteins impacting heart and brain structure.

Analysis of receptor expression in scRNAseq highlighted fibroblasts, SMCs, and monocytes/macrophages as key cellular targets of circulating proteins. Of artery-enriched receptors for cardiac IDP-associated ligands, 26/41 (63%) were expressed in aortic fibroblasts, 61% in SMCs, 56% in monocytes/macrophages, and 54% in T cells. Similarly, of artery-enriched receptors for brain IDP-associated ligands, 15/18 (83%) were expressed in aortic fibroblasts, 78% in T cells, 72% in SMCs, 44% in monocytes/macrophages (Extended Data Fig. 8a-d). These patterns were similar in carotid tissue, reinforcing the prominent roles of fibroblasts, SMCs, and monocytes/macrophages in coordinating vascular-driven signaling in both heart and brain structure.

### Pathway enrichment and protein-protein interactions

To elucidate the biological functions of imaging trait-associated proteins, we conducted pathway enrichment analyses^37^. CMR trait-associated proteins showed strong enrichment for inflammatory and immune pathways (Fig. 2h). Pathways related to blood vessel and angiogenesis also appeared but were less prominent. Fewer proteins were linked to brain IDPs, showing enrichment for pathways related to extracellular matrix (Fig. 2i). Insulin-like growth factor transport emerged as shared themes across cardiac and brain datasets (Fig. 2h,i). We also assessed protein-protein interaction (PPI) networks of IDP-associated proteins^38^. The PPI expansions for cardiac and brain IDP-associated proteins reaffirmed inflammatory, vasculature, and ECM-related pathways, respectively (Fig. 2j, k). Notably, brain IDPs-associated PPI networks were also enriched for cardiomyopathy-related pathways, reinforcing the overlap between cardiac and neurological mechanisms.

### Disease prevalence and incidence analysis

We hypothesized that plasma proteins linked to cardiac and/or brain imaging traits would be enriched for associations with cardiovascular (CVD) and neurological (NeuD) diseases. Using ICD-10 codes, we evaluated associations with 77 CVD and 67 NeuD diagnoses by logistic regression for prevalent disease and Cox regression for incident disease in 53,014 participants over a median of 14.6 years. We defined associations as clinically meaningful if the odds ratio (OR) or hazard ratio (HR) deviated from the null hypothesis by ≥20% and as statistically significant if *p*<0.05 after Bonferroni correction.

Among the 404 cardiac IDP-associated proteins, 380 (94.1%) were linked to future (incident) CVD and 345 (85.4%) to current (prevalent) CVD, substantially higher than the corresponding proportions across the entire Olink platform (43.7% and 37.8%, respectively; *p*<0.0001). Notably, most were also associated with incident (62.1%) and prevalent (53.5%) NeuD (Fig. 3a,c). Of the 76 brain IDP-associated proteins, most were associated with incident (56.6%) and prevalent (52.6%) NeuD (vs 14.9% and 20.1%, respectively for Olink overall, *p*<3×10^−18^). Remarkably, more of the brain IDP-associated proteins were associated with incident (94.7%) and prevalent (86.8%) CVD (Fig. 3b,d, Extended Data Fig. 9). The reported percentages reflect overlap between proteins associated with heart or brain IDPs and proteins associated with at least one ICD-10–defined cardiovascular or neurological endpoint, providing an aggregate measure of clinical relevance across endpoints. These overlap summaries should be interpreted as descriptive, reflecting broad concordance across clinical endpoints rather than pinpointing a single disease mechanism.

**Fig. 3.**
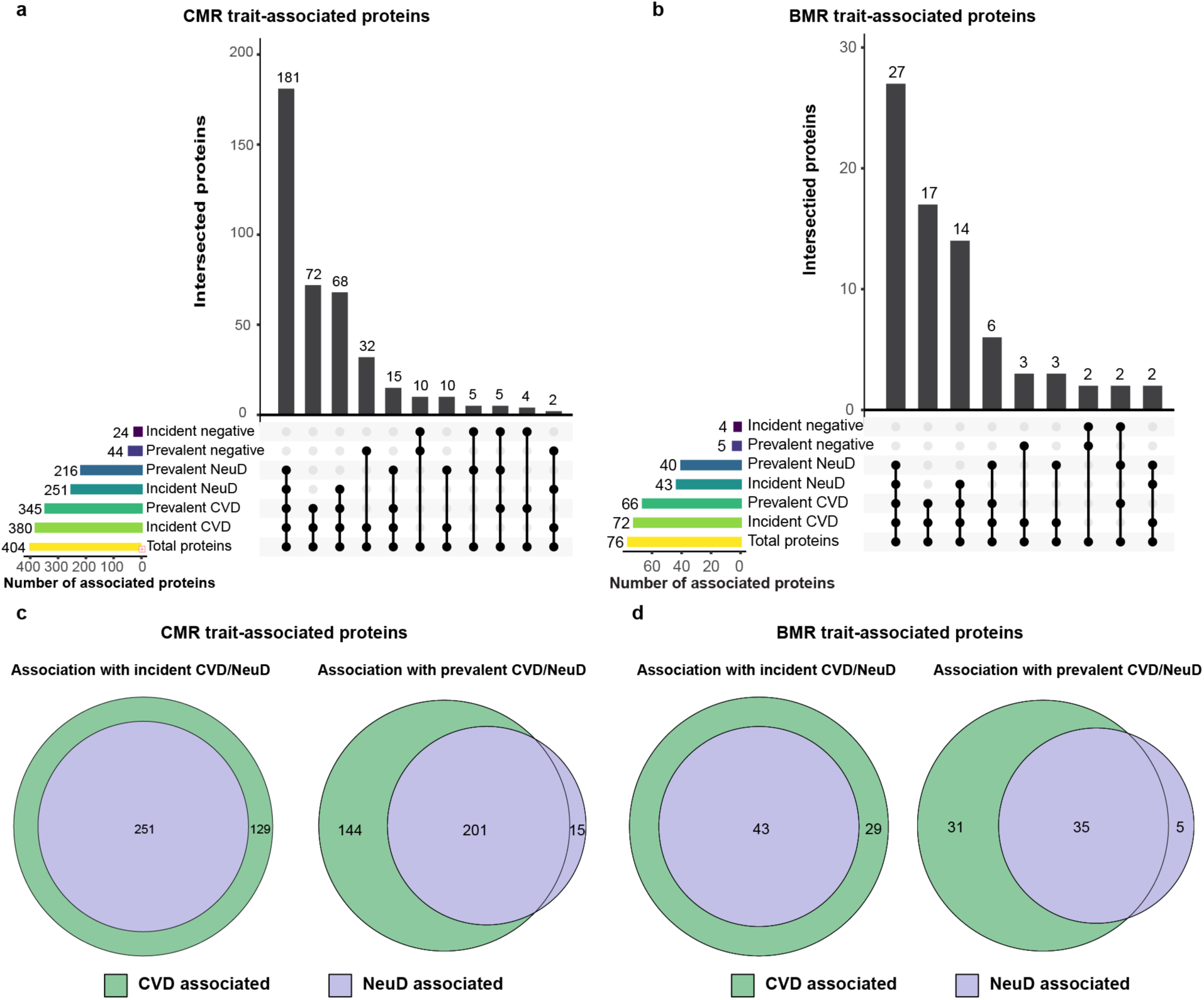
Associations between prevalent and incident disease and MRI trait-associated proteins. **a–b.** Proportions of CMR trait-associated (**a**) and BMR trait-associated (**b**) proteins linked to incident or prevalent cardiovascular disease (CVD) or neurological disease (NeuD). **c–d**. Overlap of proteins associated with incident/prevalent CVD and NeuD for CMR trait (**c**) and BMR trait (**d**) associated proteins.

Because essential hypertension (I10) is highly prevalent and could disproportionately influence composite “any CVD” summaries, we performed a sensitivity analysis excluding I10 from the cardiovascular endpoint set. The overall overlap patterns and key conclusions remained qualitatively unchanged: for prevalent CVD, overlap was complete in the full sample (CMR 345/345; BMR 66/66), with only minor sex-specific differences (male: one hypertension-only protein for both CMR and BMR; female: seven hypertension-only proteins for CMR). For incident CVD, results were identical across analyses (100% overlap), indicating that the observed CVD associations were not driven by hypertension. Stroke diagnoses were retained within the cardiovascular endpoint set to remain consistent with standard ICD-10 classification and to avoid subjective reassignment of vascular neurological events across disease categories. This is also an appropriate reflection of the shared cardiovascular–neural pathophysiology of stroke. To facilitate interpretation, all endpoint diagnoses are explicitly annotated.

Reassuringly, recognized biomarkers emerged from these analyses. NT-proBNP, routinely used in HF management, was robustly associated with prevalent HF (male OR=4.39, *p*=2.14×10^−154^; female OR=4.99, *p*=7.34×10^−50^). Galectin-3, an FDA-qualified HF risk stratification biomarker, was associated with prevalent HF (OR=1.60, *p*=1.44×10^−23^). Neurofilament light (NEFL), an established neurodegeneration marker^39,40^ was linked to incident Parkinson’s disease (HR=1.22, *p*=3.37×10^−8^). Renin, central to the RAAS, correlated strongly with prevalent HF (OR=3.86, *p*=6.2×10^−139^), consistent with successful therapies targeting RAAS in HF^41^.

Associations also emerged that underscore heart-brain interconnections. In addition to its association with HF, NT-proBNP was associated with neuropathy (mononeuropathy OR=3.95, *p*=1.7×10^−10^; polyneuropathy HR=1.71, *p*=9.05×10^−15^), and transient cerebral ischemic attacks (HR=1.24, *p*=4.1×10^−8^). While not widely appreciated, the latter two associations have been noted previously in large-scale proteomic studies^42^. Conversely, BCAN, a CNS extracellular matrix proteoglycan, was inversely associated with incident HF (HR=0.76, *p*=2.77×10^−36^) and atherosclerosis (HR=0.65, *p*=4.88×10^−18^), confirming prior proteomic findings^42,43^. GDF15 showed wide-ranging associations with CVD (HF: HR=1.65, *p*=4.99×10^−157^; atherosclerosis: HR=1.9, *p*=4.77×10^−56^) and NeuD (e.g., mononeuropathy: HR=2.90, *p*=1.54×10^−23^; cerebral infarction: HR=1.40, *p*=2.3×10^−32^). Dipeptidyl peptidase-4, an antidiabetic drug target, correlated with BMR traits (e.g., brain volume ratio) and several neurological outcomes (e.g., Parkinson’s disease: HR=0.73, *p*=1.01×10^−10^).

Some proteins had sex-specific disease associations, with eight exhibiting opposite effects in males and females. For example, collagen alpha-1(IX) chain had a positive correlation with HF in females but a negative correlation in males (e.g., HR=1.23, *p*=3.08×10^−10^ vs. HR=0.86, *p*=1.99×10^−7^), perhaps reflecting well-documented sex differences in HF phenotypes. Conversely, follitropin subunit beta (FSHb) was positively associated with incident atrial fibrillation in females (HR=1.23, *p*=2.61×10^−8^) but negatively associated in males (HR=0.85, *p*=1.55×10^−7^), potentially reflecting the increased FSHb seen in women at menopause^44^.

Most associations identified were not established or broadly recognized biomarkers. Of the 345 cardiac IDP-associated proteins linked to prevalent CVD, only 5.5% are recognized biomarkers in any organ system according to MarkerDB^45^. Similarly, 5.6% of the 216 cardiac IDP-associated proteins linked to prevalent NeuD are known biomarkers. Brain IDP-associated proteins linked to disease showed similar patterns. Thus, the large majority of protein-disease associations identified implicate new candidate biomarkers in CVD and/or NeuD.

We further performed sensitivity analyses incorporating more extensive confounder adjustment, including socioeconomic variables (Townsend deprivation index and educational attainment), cardiometabolic risk factors (systolic blood pressure, dyslipidemia, and type 2 diabetes), and lifestyle factors (smoking and alcohol consumption). The results showed substantial consistency with the primary analyses. For prevalent CVD, 326 of 345 CMR IDP-associated proteins and 65 of 66 BMR IDP-associated proteins remained significant after additional adjustment. Similarly, for prevalent neurologicalf disease, 182 of 216 CMR IDP-associated proteins and 36 of 40 BMR IDP-associated proteins remained significant. Comparable robustness was observed for incident outcomes: 366 of 380 CMR IDP-associated proteins and 68 of 72 BMR IDP-associated proteins remained significant for incident CVD, and 204 of 251 CMR IDP-associated proteins and 35 of 43 BMR IDP-associated proteins remained significant for incident neurological disease.

### Genetic association analysis: implications for causal links

To determine whether the proteins identified could play causal roles in disease, we performed MR using genome-wide significant *cis*-pQTLs (instrumental variants, IVs) for proteins linked to imaging traits and 75 cardiovascular (CVD) and neurological/psychiatric (NPDs) outcomes. Many of these *cis*-pQTLs are reported risk variants for relevant conditions in GWAS^46^, consistent with possible etiological roles. Outcomes were grouped into five CVD categories: coronary artery disease (CAD), HF, arrhythmia, hypertension, and stroke, and three NPDs categories: psychiatric disorders, neurodegenerative diseases, and dementia. We utilized a two-sample MR framework with 12 methods to account for pleiotropy and heterogeneity, complemented by Steiger directionality testing to confirm exposure-to-outcome effects. To minimize bias from genetic overlap, disease GWAS summary statistics were sourced from multiple repositories, including the NHGRI-EBI GWAS Catalog^46^, MRC IEU OpenGWAS^29,30^, and FinnGen^47^.

Among 367 proteins linked solely to CMR traits, 327 had valid autosomal *cis*-pQTLs for MR. Of these, 225 (68.8%) displayed potential causal relationships with at least one disease outcome at a 5% FDR: 35.5% with both CVD and NPDs; 17.1% only with CVD; 16.2% only with NPDs. Psychiatric disorders, neurodegenerative diseases, stroke, and CAD were all commonly implicated (∼26%-31%) (Fig. 4a,d). Of 39 proteins linked solely to BMR traits, 36 had valid *cis*-pQTLs. Twenty-five (69.4%) demonstrated potential causal associations: 41.7% with both CVD and NPDs; 19.4% only with CVD; 8.3% only with NPDs. Stroke (36.1%), neurodegenerative diseases (33.3%), and psychiatric disorders (30.6%) were commonly implicated with CAD and HF, slightly lower at 19% each (Fig. 4b,d). Among 37 proteins associated with both CMR and BMR traits, 35 had valid *cis*-pQTLs. Twenty-eight (80%) showed potential causal links to CVD and/or NPDs: 45.7% in both organ systems: 14.3% solely in CVD; 20% solely in NPDs. In a pattern similar to CMR IDP-associated proteins, psychiatric disorders (45.7%), neurodegenerative diseases (42.9%), stroke (37.1%), and CAD (34%) were the most commonly linked outcomes (Fig. 4c,d).

**Fig. 4.**
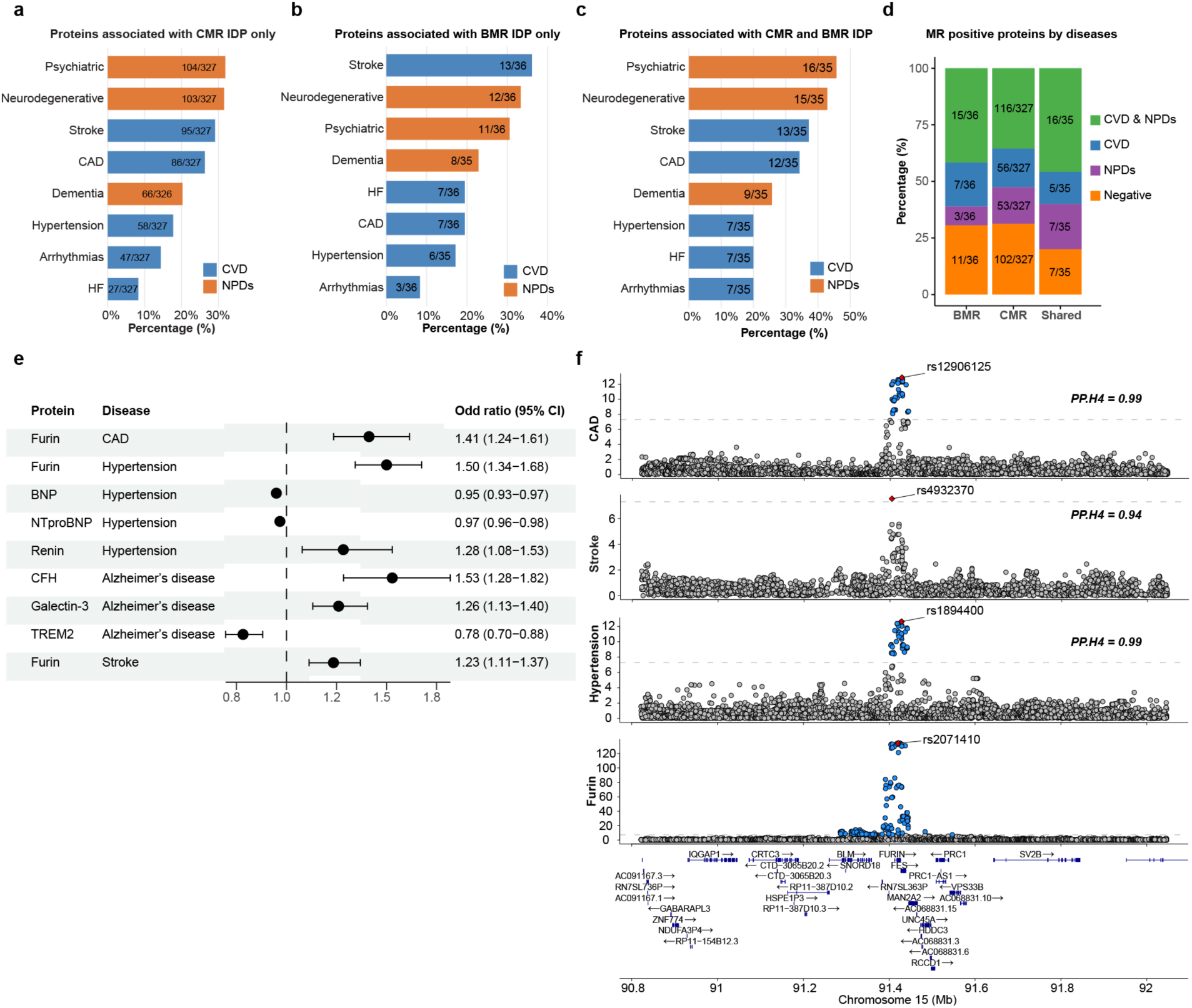
Genetic causal inference between MRI-associated proteins and CVD/neurological-psychiatric diseases (NPDs). **a–c.** Proportion of proteins showing significant Mendelian randomization (MR) evidence for CVD or NPDs among CMR IDP-only (**a**), BMR IDP-only (**b**), or both CMR and BMR IDP (**c**) associated proteins. **d**. Overall percentage of proteins significantly associated with CVD or NPDs by MR analysis. **e**. Distribution of MR estimates for robust protein-disease pairs. **f**. Genetic colocalization of Furin with hypertension, stroke, and coronary artery disease (CAD) on chromosome 15. Genes in the locus are shown along the x-axis; the y-axis displays −log_10_(*p*). Blue points represent trait-associated variants; highlighted variants indicate trait-specific lead variants. HF, heart failure; CFH, complement factor H; TREM2, triggering receptor expressed on myeloid cells 2.

Our analyses confirmed multiple known causal roles. For example, Renin and BNP, both known to regulate blood pressure, were both causally linked to hypertension (*p*=4.97×10^−3^ and 1.40×10^−9^, respectively). Triggering receptor expressed on myeloid cells 2 (TREM2), one of the genetic risk factors for late-onset Alzheimer’s disease (AD), was causally associated with AD (OR=0.78, *p*=4.57×10^−5^). However, there were also false negatives. Proteins including PCSK9, Angiotensin-converting enzyme (ACE), and Apolipoprotein E (APOE) did not survive Bonferroni correction for IDP association. Thus, the stringent criteria applied to enhance confidence in candidates identified also eliminated some proteins of interest.

Importantly, most of the potentially causal protein-disease associations identified were novel, with only 11.9% current drug targets^48^. Furin, a proprotein convertase correlated with ventricular end-diastolic volumes, showed possible causal roles in hypertension (OR=1.50, *p*=6.56×10^−13^), CAD (OR=1.41, *p*=4.47×10^−7^), and stroke (OR=1.23, *p*=6.66×10^−5^), consistent with the recognition that its targets regulate cholesterol metabolism, inflammation, and blood pressure^49,50^. Many novel candidates identified are linked to diseases for which therapies are limited, such as neurodegenerative disorders including AD, Parkinson’s disease, and dementia. Galectin-3, a recognized biomarker for HF^51^, was causally linked with Parkinson’s disease (OR=1.26, *p*=2.85×10^−5^). Complement Factor H (CFH), a key regulator of complement-mediated immune responses, was inversely correlated with multiple LV phenotypes and showed a causal association with dementia in AD (OR=1.53, *p*=1.68×10^−6^), highlighting the connection between cardiac remodeling and neurodegeneration (Fig. 4e).

Although the primary MR analyses incorporated outcome GWAS from multiple independent resources to minimize bias, we further validated and replicated the findings using two additional large-scale plasma proteomics datasets from deCODE^52^ and Fenland^53^, both generated using the SomaScan platform. Among UKB plasma proteins significantly associated with CMR and/or BMR imaging traits, 207 and 184 were also profiled in the deCODE and Fenland datasets, respectively. Applying the same MR analysis pipeline, we observed high replication rates. In the comparison between UKB and Fenland proteomics, 77% (97 of 126 proteins) showed consistent and significant causal associations in both datasets. Of these, 72% (62 proteins) were associated with CVD outcomes and 63% (62 proteins) with NPDs outcomes. Similarly, between UKB and deCODE proteomics, 70% (101 of 144 proteins) were significant in both datasets, with 57% (55 proteins) and 57% (65 proteins) associated with CVD and NPD outcomes, respectively.

We further used Bayesian colocalization to determine whether the same variant influenced both protein levels and disease risk. Among CMR IDP-specific proteins, 161 of 313 MR-positive signals (51.4%) showed moderate-to-high colocalization, particularly for CAD, hypertension, and stroke. Similar trends were observed for BMR IDP-specific and shared-protein signals, including near-complete colocalization of the CELSR2 locus with CAD (*PP.H4*≈1) and robust colocalization of Furin with hypertension, CAD, and stroke (*PP.H4* = 0.94-0.99) (Fig. 4f). Although colocalization strengthens causal inference, the absence of colocalization does not rule it out^54^; complex loci may harbor multiple independent signals. Nonetheless, these results indicate that a substantial fraction of IDP-associated proteins display robust genetic evidence for causal effects in CVD and NPDs and warrant further investigation as therapeutic targets.

## DISCUSSION

In this study, we integrated large-scale plasma proteomics, high-resolution cardiac and brain MRI data, genetic information, and clinical outcomes from the UKB to investigate the role of plasma proteins in connecting cardiovascular and neurological health. Our integrative genomic and proteomic analyses substantially expand current understanding of the genetic and molecular underpinnings of heart-brain interactions. We identified hundreds of circulating proteins associated with imaging traits in both the heart and brain. Most of these proteins also predicted future CVD and NeuD, underscoring their potential utility as clinical biomarkers. Genetic analyses utilizing MR and genetic colocalization provided robust evidence supporting potentially causal roles for many of these proteins, suggesting they may be candidates for therapeutic intervention. In both cases, the large majority of these associations are novel, providing an atlas of hundreds of new candidate biomarkers and therapeutic targets for future investigation (Fig.5).

**Fig. 5.**
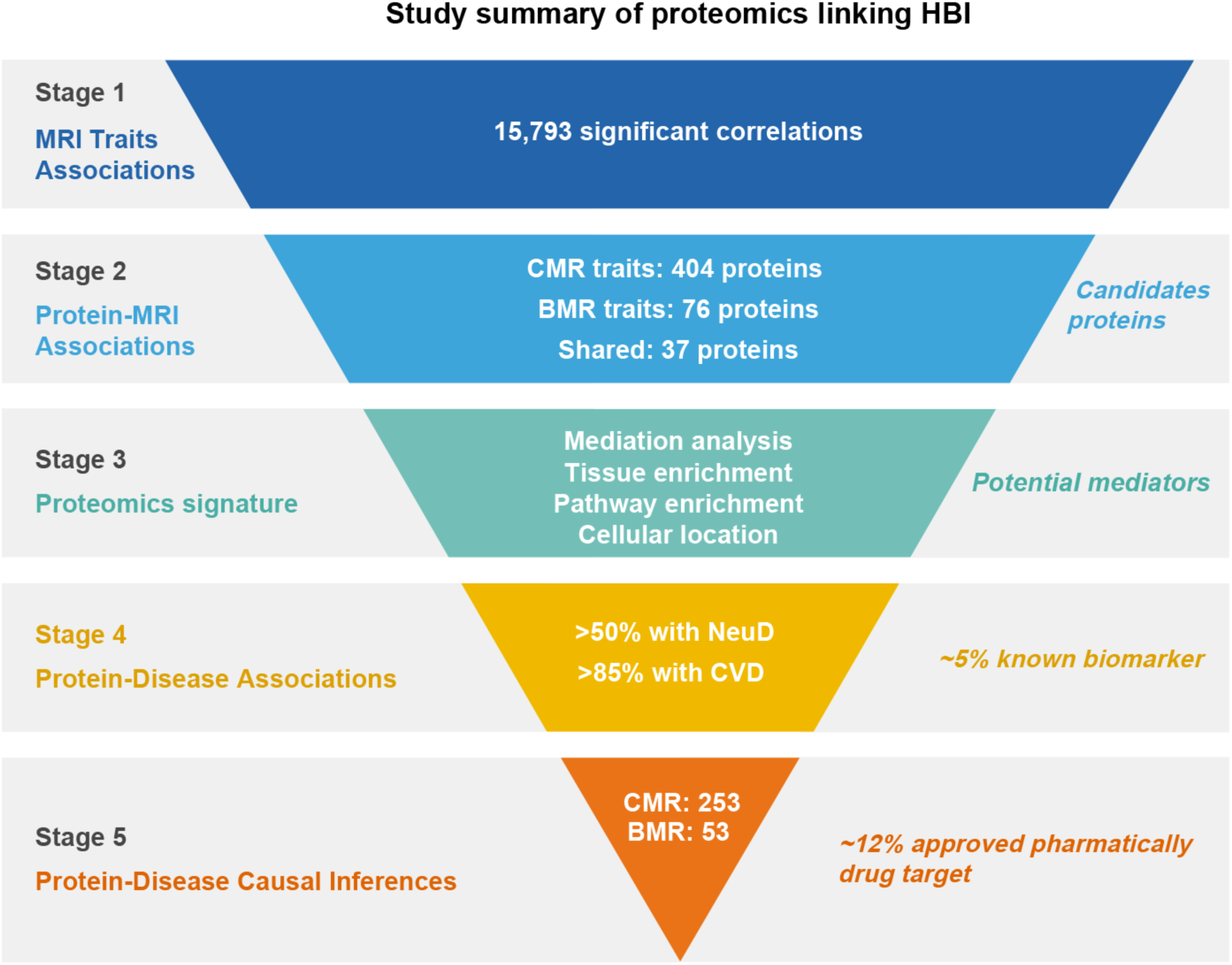
Summary of the study. Graphical overview of the heart-brain interaction (HBI) framework investigated in this study, illustrating how MRI traits, circulating proteins, and disease outcomes inform one another. CMR, cardiac MRI IDP; BMR, brain MRI IDP; CVD, cardiovascular diseases; NeuD, neurological diseases.

Our findings advance the understanding of heart-brain connections in multiple important ways. First, they underscore the value of IDPs as endophenotypes that provide novel insight into disease associations and mechanisms. The large majority of proteins correlated with imaging traits had strong associations with cardiovascular and/or neurological disease. Among the cardiac IDP-associated proteins, 85.4% were associated with existing (prevalent) and 94.1% with new (incident) disease in one or both of these organ systems. For brain IDP-associated proteins, 86.8% were associated with prevalent and 94.7% with incident disease in at least one of these organ systems. The close connection between structure and function for both the heart and brain likely underlies the striking enrichment of IDP correlates for indicators of existing and future disease in these organs. Whether a similar strategy would yield comparable results for other organs is worthy of future investigation.

Remarkably, proteins associated with IDPs in one organ were strongly associated with disease not only in that organ but in the other as well. For cardiac IDP-associated proteins, most were associated with prevalent (53.5%) or incident (62.1%) neurological disease, though these were fewer than the 85.4% and 94.1%, respectively, associated with cardiovascular disease. For brain IDP-associated proteins, the association was stronger for cardiovascular than neurological disease. 52.6% and 56.6 were associated with prevalent and incident neurological disease, respectively, whereas an extraordinary 86.8% and 94.7% were associated with prevalent and incident cardiovascular disease. There was also notable overlap in these protein sets (Fig.3). For example, *all* of the cardiac IDP-associated proteins that were predictive of future neurological disease also predicted future cardiovascular disease, and *all* of the brain IDP-associated proteins that predicted future neurological disease also predicted cardiovascular disease. Together these data reinforce the close connections between heart and brain health and the importance of common circulating proteins in both organs. Recent evidence suggests that acute organ injury (e.g., stroke) can induce long-lasting immune and epigenetic remodeling that alters circulating protein profiles^55^. Such effects may plausibly contribute to observed disease associations, supporting protein-mediated links between cardiovascular and neurologic disease, while also raising the possibility that some circulating signals reflect prior or comorbid conditions rather than the primary organ process of interest. Although our incident analyses reduce bias related to baseline disease status, future studies incorporating more refined disease groupings (e.g., hard CVD events, stroke subtypes, and dementia subtypes), adjudicated event timing, longitudinal proteomic profiling, and immune and epigenetic measurements will be essential to further clarify the mechanistic interpretation of these proteome-heart-brain relationships.

In many instances, circulating proteins may associate with heart and brain health independently. However, we also found statistical evidence that some proteins may transmit effects from one organ to the other. Among 17 proteins associated with paired traits amenable to mediation analysis, 10 proteins mediated significant relationships from brain to heart imaging traits, and nine mediated relationships from heart to brain traits (Extended Data Fig. 3c,d). While mediation analysis cannot prove causality, these analyses suggest circulating proteins may act as conduits for heart-brain signaling. Notably, GDF15 emerged as a dominant candidate, mediating 22 of 27 structural associations between heart and brain, with higher GDF15 levels generally correlating with smaller brain volumes. In a murine model, cardiac-specific overexpression of GDF15 produced reductions in both heart and brain weights and volumes, mirroring the human imaging associations. This experimental evidence aligns with and strengthens the population-based inferences, lending biological plausibility to the role of GDF15 as a molecular mediator of heart-brain interactions. Although our findings support a mechanistic role for GDF15 in heart–brain communication, the therapeutic value of modulating this pathway remains uncertain and is likely context- and disease-stage–dependent, requiring rigorous prospective clinical testing to assess both safety and efficacy.

The tissues most enriched for proteins associated with brain and/or cardiac IDPs were arteries, the brain, and endocrine organs, while the heart was a modest contributor (Fig. 2). Based on scRNAseq data, artery-enriched proteins were predominantly expressed in fibroblasts, smooth muscle cells, and macrophages, emphasizing the contribution of these cellular components to circulating proteins. Given the critical dependence of the heart and brain on arterial blood supply, the prominence of artery-enriched proteins is understandable. Pathway analyses revealed that proteins linked to cardiac traits frequently mapped to inflammatory and cytokine-related processes, whereas those associated with brain traits were more commonly involved in ECM remodeling. We were surprised, however, by the prominence of brain-enriched proteins and modest representation of heart-enriched proteins, particularly considering the BBB reduces levels of brain-derived proteins in the plasma. Of course, these analyses are only suggestive of the likely protein sources and would need to be verified experimentally. Tissue and cell-type localization inferred from GTEx and/or reference single-cell datasets should be interpreted with caution, as expression patterns in healthy or baseline tissue may differ substantially in disease due to altered cell states and cellular composition. Accordingly, these results are intended to provide contextual, hypothesis-generating insight into potential protein sources rather than definitive attribution, and will require independent validation in disease-relevant cohorts and experimental models. Nevertheless, they suggest an asymmetry in which proteins relevant to heart and brain structure (and disease) are more likely to be enriched in (and thus arise from) the brain than the heart, and underscore the overarching importance of the vasculature in both organs. They also highlight the role of inflammation and extracellular matrix in heart and brain health.

From a clinical perspective, the accessibility of plasma proteins makes them attractive candidates as diagnostic or prognostic biomarkers and genetically validated therapeutic targets. Although some of the disease-related proteins identified are already recognized clinical biomarkers (e.g., NT-proBNP for HF, Renin for hypertension)^42,43^, over 90% of them have not yet been established or validated for clinical use. This observation not only contextualizes the confirmatory aspects of the study but also underscores its contribution in expanding the landscape of previously underrecognized protein–disease associations deserving of further investigation. To determine which of these might lie in the pathogenic pathway and be potential therapeutic targets, we used MR and genetic colocalization. This approach confirmed established causal relationships (e.g., Renin and BNP in hypertension). However, most proteins for which MR was consistent with a causal role in CVD or NeuD were not previously known to play these roles. In many cases, these proteins were implicated in conditions for which we currently lack highly effective treatments (e.g., CFH in Alzheimer’s disease). Approximately 85% of the proteins identified as potentially causal do not currently have approved pharmacotherapies. However, if validated as therapeutic targets, plasma proteins would be tractable targets for therapeutic strategies, including targeted antibodies or antisense, with a high likelihood of success.

Several limitations warrant consideration. First, region-specific CMR-BMR IDP associations should be interpreted with caution, as they may reflect differential sensitivity or reliability of specific imaging features rather than anatomically localized mechanisms. We therefore treat them primarily as quantitative anchors for downstream protein-based analyses of heart-brain interactions. Second, the UKB population is predominantly of European ancestry, limiting generalizability. Although we included race as a covariate in our models, validation in more diverse cohorts will be important. Third, the available proteomic data were limited to 2,922 proteins measured on the Olink platform and reported as normalized protein expression (NPX) values rather than absolute concentrations. As a result, relevant proteins may be missing, and effect sizes are relative, suggesting that our analyses likely underestimate the contribution of circulating proteins to heart–brain disease mechanisms. Fourth, the Bonferroni correction applied to imaging-derived phenotype associations is highly stringent and likely increases false negative rates. Indeed, several well-established disease-related proteins (e.g., APOE, PCSK9, and ACE) did not withstand this threshold and others (e.g., troponin I and apolipoprotein B) lacked suitable *cis*-pQTLs required for Mendelian randomization. Given our goal of identifying high-confidence candidates as new biomarkers or therapeutic targets for investigation, these conservative analytic choices were intentional, albeit at the cost of underestimating the full spectrum of biologically relevant proteins. Finally, we want to emphasize that the novel disease associations and genetically supported mechanistic pathways identified here need to be validated in more experimental and prospective clinical studies before being considered actionable.

Despite these limitations, our findings offer a comprehensive, integrated atlas of plasma proteins linking cardiovascular and neurological structure and disease. The striking overlap among these proteins underscores the intimate interconnections between these organs. These findings provide a map for future validation of new biomarkers and genetically therapeutic targets in heart and brain disease.

## METHODS

### Ethical approval

The UKB operates under ethical approval from the North West Multi-centre Research Ethics Committee as a Research Tissue Bank (RTB). All participants provided written informed consent, and no additional approvals were required under the RTB framework. All mice were maintained and studied using protocols in accordance with the NIH Guide for the Care and Use of Laboratory Animals and approved by MGH Animal Care and Use Committees (protocol number 2015N000029) or by the Institutional Animal Care and Use Committee (IACUC) of Beth Israel Deaconess Medical Center (protocol number 072-2020).

### Animal studies

Wild type 12-week-old male mice were obtained from The Jackson Laboratory (C57BL/6J, #000664). Mice were fed a rodent chow diet with 12-hour light and dark cycles. Animal related study was performed at the Massachusetts General Hospital Cardiovascular Research Center.

### Adeno-associated virus

Recombinant adeno-associated virus serotype 9 (AAV9) vectors were cloned and propagated by VectorBuilder (VectorBuilder Inc, Chicago, USA). Vector plasmids utilized are the following: AAV9-cTnT-GFP - VB240220-1278nja, AAV9-cTnT-mGdf15 - VB240220-1208uec. Recombinant AAV was produced in HEK293T cells. AAVs were administered at a dose of 5×10^11^ viral genomes/mouse by diluting in a final volume of 200 µl PBS via tail-vein injection using 1 ml 30 Ga insulin syringes (B.H. Supplies). Mice were maintained in the sedentary state for 4 weeks following AAV administration, after which they were individually housed subjected to voluntary wheel running using in-cage 4.5-inch diameter voluntary running wheels in individual cages (Starr Life Sciences Corp) for 7 days before necropsy and tissue analysis.

### Quantitative real-time PCR for mRNA expression

Total RNA from heart was extracted using TRIzol (Invitrogen), purified with RNeasy Mini spin columns (Qiagen), and reverse transcribed using a HighCapacity cDNA Reverse Transcription kit (Applied Biosystems). The resulting cDNA was analyzed by RT-qPCR using SYBR green, fluorescent dye 2×qPCR master mix (Promega) in a QuantStudio 6 Flex Real-Time PCR System (Applied Biosystems). The Rplp0 mRNA was used as a loading control, and fold change was calculated using the ΔΔCt method.

### Plasma ELISA measurement

Plasma samples were collected from mice injected with either AAV9-cTnT-Gdf15 or AAV9-cTnT-GFP control vectors. GDF15 concentrations were quantified using a commercially available mouse GDF15 enzyme-linked immunosorbent assay (ELISA) kit (DY6385, R&D Systems) following the manufacturer’s instructions. All samples were measured in duplicate, and absorbance was read at 450 nm using a microplate reader. Standard curves were generated using provided calibrators, and plasma GDF15 concentrations were calculated accordingly.

### Heart/brain weight and volume measurements

At the study endpoint, mice were euthanized, and heart and brain tissues were carefully excised, blotted dry, and weighed to the nearest milligram. Organ weights were normalized to tibia length to account for body size differences. Brain volume was determined by water displacement, wherein the freshly excised brain was fully submerged in a graduated cylinder containing distilled water, and the volume of displaced water was recorded as the brain volume (Archimedes’ principle). All measurements were performed in a blinded manner to treatment group to minimize bias.

### Population study design

This study utilized data from the UK Biobank (UKB), a large-scale biomedical resource comprising more than 500,000 participants aged 40-69 years at baseline (2006-2010), followed up at several subsequent visits. The UKB includes genome-wide genotyping data, magnetic resonance imaging (MRI), electronic health records, blood and urine biomarkers, and physical/anthropometric measurements. Cardiac (CMR) and brain (BMR) MRI scans were acquired during the later assessment visits. The present analysis focused on participants who underwent CMR and BMR imaging and had available plasma proteomics data.

### Cardiac and brain MRI imaging-derived phenotypes

CMR imaging-derived phenotypes (IDPs) were extracted from raw CMR scans using the established methodology, as previously described^56^. The UKB released 82 quantitative CMR metrics grouped into six categories: left ventricular, left atrial, right ventricular, right atrial, ascending aorta, and descending aorta. These metrics provide structural and functional information on cardiac morphology and performance. In total, 39,676 participants underwent CMR scanning during the first imaging assessment revisit.

BMR IDPs were obtained from raw images acquired through five primary MRI modalities: 1) T1-weighted imaging (T1), highlighting brain volumes and cortical thickness; 2) T2-weighted Fluid Attenuation Inversion Recovery (FLAIR), highlighting pathological changes (e.g., white matter hyperintensities); 3) Susceptibility-weighted imaging (SWI), detecting tissue susceptibility variations (e.g., venous structures, microbleeds); 4) Diffusion-weighted imaging (DWI), capturing white matter microstructure and connectivity; 5) Resting-state fMRI (rfMRI), measuring blood-oxygen-level-dependent fluctuations for regional connectivity and functional activity. Among the DWI data, diffusion MRI weighted means provide overall diffusion properties, while tract-based analyses (probabilistic tractography, DTI/NODDI) focus on microstructural integrity in distinct white matter pathways. Skeletonized diffusion measurements further concentrate on the central core of white matter tracts, enabling sensitive detection of subtle changes in microstructure. Task-fMRI activation IDPs were excluded due to low reproducibility and heritability, as previously reported^21^.

The UKB processed these modalities using established pipelines, generating 3,915 IDPs for 47,453 participants, categorized into 11 groups (e.g., global/regional brain volumes, white matter hyperintensity, cortical thickness/surface area, SWI-derived metrics, DTI/NODDI parameters, rfMRI component amplitudes, functional connectivity metrics). Replicated cortical IDPs from multiple parcellation schemes (e.g., Desikan-Killiany, DKT) were included to ensure robustness. The March 2024 data release served as the source for BMR IDPs^57–61^.

### Plasma proteomics data

Plasma proteomic measurements were sourced from the UK Biobank Pharma Proteomics Project (UKB-PPP)^20^, which used the Olink Explore 3072 proximity extension assay (PEA), an antibody-based technique. Between April 2021 and February 2022, blood plasma samples from 53,014 participants were analyzed, yielding 2,941 protein analytes corresponding to 2,923 unique proteins. The released dataset (as of September 2023) provides normalized protein expression (NPX) values across panels related to inflammation, oncology, cardiometabolic processes, and neurological functions. Detailed documentation of the Olink proteomics data is available through the UK Biobank showcase platform (https://biobank.ndph.ox.ac.uk/showcase/refer.cgi?id=4654).

### Data processing

We applied extensive quality control (QC) measures to MRI IDPs and proteomics data. For CMR and BMR IDPs, both manual inspection and automated QC protocols were used. IDPs were excluded if they were incomplete or displayed artifacts related to head motion, anatomical abnormalities, or other technical issues. Individuals lacking complete T2-FLAIR and T1 data in FreeSurfer analyses were also removed. The Olink proteomics data underwent QC by the UKB-PPP team prior to release^20^.

We excluded outliers in IDPs exceeding eight times the median absolute deviation from the median. Non-numeric columns, variables with over 90% missingness, and those having >95% identical values were removed. Highly correlated IDPs or proteins were also excluded to prevent redundancy. For instance, Glioma Pathogenesis-Related Protein 1 was removed due to >90% missing data. Low-variability measures (e.g., 5th ventricle volume, non-white matter hypo-intensities) were similarly discarded.

Given the inherent noise in functional connectivity estimates for individual region-pairs^59^, the 1,695 rfMRI connectivity IDPs were dimension-reduced into six summary IDPs using data-driven feature extraction^59^. These were treated as composite IDPs representing linear combinations of the original connectivity features. More details on this process can be found on the FMRIB UK Biobank Resource webpage (http://www.fmrib.ox.ac.uk/ukbiobank/).

After QC, 39,676 participants remained with ≥1 of 82 CMR IDPs, 47,453 participants with ≥1 of 2,222 BMR IDPs, and 53,014 participants with 2,922 NPX proteomic measurements. IDPs and proteomic NPX values were rank-based inverse Gaussian transformed for approximate normality.

To reduce spurious associations arisng from confounding, we applied a two-stage residualization framework^62^ throughout the study. In the first stage, imaging-derived phenotypes (IDPs) and non-imaging variables were residualized separately with respect to prespecified confounders using linear regression. This deconfounding step removed the effects of known demographic and technical factors and generated residualized values for each variable. In the second stage, associations of interest (e.g., correlations or regression analyses) were evaluated between these residualized variables.

Across all analyses, established covariates were regressed out from IDPs, cognitive measures, and proteomic data. In the primary exploratory analyses, covariates included age, sex, body mass index (BMI), height, weight, ethnicity (White British vs. other), and modality-specific technical factors. For cardiac MRI (CMR), technical covariates included heart rate and imaging site^19,56^. For brain MRI (BMR), technical covariates included head size, imaging site, head motion during resting-state and diffusion MRI, head position, and scanner table position^57,58,60,61^. For Olink proteomics, technical covariates included assay batch and assessment center. In sensitivity analyses, we additionally adjusted for socioeconomic variables (Townsend deprivation index and educational attainment), lifestyle factors (smoking status and alcohol consumption), and cardiometabolic risk factors (systolic blood pressure, dyslipidemia, and type 2 diabetes status). Extreme values in covariates (e.g., scanner table position or head motion) were retained and transformed using rank-based inverse normal transformation where appropriate. The resulting deconfounded and normalized residuals served as the basis for all downstream analyses.

### Correlation between cardiac and brain MRI-derived phenotypes

A univariate, pairwise Pearson correlation was performed between 82 CMR IDPs and 2,222 BMR IDPs in 32,859 individuals with measurements for both organs. All IDPs had undergone the aforementioned QC, normalization, and de-confounding. The confounders included age, sex, BMI, height, weight, ethnicity, heart rate, imaging site, head size, head motion during resting-state and diffusion MRI, head position, and scanner table position. Pearson’s correlation coefficients were calculated, and degrees of freedom varied according to data completeness. After multiple comparison correction (Bonferroni), the significance threshold was set at *p*<2.74×10^−7^ (equivalent to a family-wise error rate of 0.05 across 182,204 tests).

### Correlation between brain MRI-derived phenotypes and cognition function

A univariate, pairwise Pearson correlation was performed between eight cognition assess function and 2,222 BMR IDPs in 36,349 individuals. Cognition assessment included numeric memory test (maximum digits remembered correctly), prospective memory test (number of attempts, final attempt correct, prospective memory result: Instruction not recalled, either skipped or incorrect/correct recall on first attempt/ correct recall on second attempt), and Fluid intelligence score (number of fluid intelligence questions attempted within time limit). All IDPs had undergone the aforementioned QC, normalization, and de-confounding. Confounders include age, sex, BMI, ethnicity, head size, imaging site, head motion during resting-state and diffusion MRI, head position, scanner table position, townsend deprivation index, educational attainment, smoking status, alcohol consumption, systolic blood pressure, dyslipidemia, and type 2 diabetes. Pearson’s correlation coefficients were calculated, and degrees of freedom varied according to data completeness. After multiple comparison correction (Bonferroni), the significance threshold was set at *p*<2.81×10^−6^ (equivalent to a family-wise error rate of 0.05 across 17,776 tests).

### Correlation between proteomics and MRI-derived phenotypes

Parallel correlation analyses were performed between plasma proteomics (Olink NPX) and CMR/BMR IDPs. Quantile-normalized, de-confounded proteomic values from the first profiling session were then correlated with CMR IDPs (4,966 participants; 82 IDPs) and BMR IDPs (5,686 participants; 2,222 IDPs). The primary analysis confounders include age, sex, BMI, height, weight, ethnicity, heart rate, imaging site, head size, head motion during resting-state and diffusion MRI, head position, scanner table position, Olink proteomics assay batch and assessment center. In sensitivity analyses, we additionally adjusted for Townsend deprivation index, educational attainment, smoking status, alcohol consumption, systolic blood pressure, dyslipidemia, and type 2 diabetes status. Pearson’s correlation coefficients were computed for 2,922 proteins, yielding 239,604 pairwise tests for CMR IDPs and 6,492,684 for BMR IDPs. Bonferroni-corrected thresholds were set at *p*<2.10×10^−7^ for CMR and *p*<7.70×10^−9^ for BMR to maintain a family-wise error rate of 0.05.

### Mediation analysis

To investigate whether specific proteins mediate heart-brain interactions identified via CMR-BMR and proteomics correlations, we conducted linear regression-based mediation analyses^63^. We first identified 37 plasma proteins associated with both CMR and BMR IDPs and further restricted analyses to 17 proteins that were also significantly associated with the corresponding CMR-BMR IDP pairs (21 CMR IDPs and 72 BMR IDPs). All imaging traits and protein levels were inverse rank-based normalized and standardized before analysis.

For each CMR-BMR-protein triplet, three models were fitted: (i) a total-effect model regressing the dependent IDP on the independent IDP; (ii) an exposure-mediator model regressing protein levels on the independent IDP; and (iii) a direct-effect model regressing the dependent IDP on both the independent IDP and the mediator protein. Ordinary least squares regression was used throughout. The indirect (mediated) effect was estimated as the product of the exposure-mediator and mediator-outcome coefficients and its significance was assessed using non-parametric bootstrapping (2,000 resamples; α = 0.0005).

Models were adjusted for age, sex, BMI, height, weight, heart rate, head size, imaging site, head motion during rfMRI/dMRI, brain position, scanner table position, Olink assay batch, Ethnicity. Multiple testing was controlled using the Benjamini-Hochberg false discovery rate (FDR<0.05). Mediation was considered present when both the total and indirect effects were statistically significant and directionally consistent.

### Tissue and cell type enrichment analysis

We performed tissue enrichment analyses using the enhancing Genotype-Tissue Expression (eGTEx)^31,32^ data covering 32 tissues. Proteins were classified as tissue-specific (score ≥4 in one tissue, <2.5 in others), tissue-enriched (score ≥2.5 in at least one tissue, but not specific), broadly expressed (score <2 in all tissues), or unknown. For simplicity, these 32 tissues were consolidated into 15 broader categories (e.g., “Heart,” “Brain,” “Artery”). Arterial tissues (aorta, coronary, tibial) were grouped together; brain cerebellum/cortex were grouped as “Brain,” etc..

We then examined ligand-receptor interactions via the Omnipath database^35^ by mapping plasma proteins to transmitter/ligand roles and restricting receptors to those annotated as ion channels, transporters, or membrane-bound receptors. Cell-type specificity in arterial tissues was further investigated using single-cell RNA-seq (scRNA-seq) data for the aorta (GSE155468)^33^ and carotid (GSE253903)^34^, processed in R (Seurat v5) with standard quality filters. Cells were classified into fibroblasts, smooth muscle cells, macrophages/monocytes, T cells, endothelial cells, NK cells, and other immune cells based on known markers. Dimensionality reduction (UMAP) and unsupervised clustering (FindCluster in Seurat) guided cell-type annotation, and differentially expressed genes were identified (log_2_ fold change > 0.25, expressed in ≥25% of cells) to confirm cell-type enrichment.

### Biological process and pathway enrichment

Biological processes and pathways were assessed using clusterProfiler^37^, drawing on GO Biological Processes, KEGG, and REACTOME. We applied Benjamini-Hochberg FDR (<0.05) for multiple comparisons. Because proteins often function in clusters, we integrated protein-protein interaction (PPI) data from STRING^38^ (version 12.0), restricting to high-confidence interactions (score > 0.7) from curated databases and experimental sources. For the 76 BMR IDP-associated proteins and 404 CMR IDP-associated proteins, we incorporated their immediate functional neighbors (first-degree interactions) at a 12.5% extension rate. Enrichment analyses were then repeated on these augmented protein sets.

### Disease prevalence and incidence analysis

We evaluated whether CMR IDP and BMR IDP-associated proteins were linked to prevalent or incident disease using 77 cardiovascular (ICD-10: I00-I99) and 67 neurological (ICD-10: G00-G99) conditions. We examined a broad set of ICD-10–defined cardiovascular (I-codes) and neurological (G-codes) outcomes to generate hypotheses linking IDP-associated proteins to clinically recorded disease endpoints in the UKB-PPP subset. Disease definitions incorporated primary care records, inpatient data, death registries, and self-reported histories, all mapped to three-character ICD-10 codes. We recognize that ICD-10 codes differ in diagnostic specificity and positive predictive value across conditions; therefore, these endpoint analyses are intended to complement the imaging and proteomic associations rather than serve as adjudicated clinical outcomes.

Prevalent disease was defined as having been diagnosed on or before the date of blood collection, while incident disease referred to new diagnoses thereafter. Logistic regression analyzed prevalence, and Cox regression (with September 2023 as the censoring date) analyzed incidence, age used as underlying timescale in the time-to-event analysis The primary analysis confounders include age, sex, BMI, height, weight, ethnicity, heart rate, imaging site, head size, head motion during resting-state and diffusion MRI, head position, scanner table position, Olink proteomics assay batch and assessment center. In sensitivity analyses, we additionally adjusted for Townsend deprivation index, educational attainment, smoking status, alcohol consumption, systolic blood pressure, dyslipidemia, and Type 2 diabetes status. A Bonferroni correction (*α* = 0.05) determined significance. We defined associations as clinically meaningful if the odds ratio or hazard ratio deviated from the null hypothesis by ≥20% and as statistically significant if *p*<0.05 after Bonferroni correction. For transparency and interpretability, we report case counts (prevalence and/or incident events) for each ICD-10 endpoint.

### Mendelian randomization analysis

We used a two-sample Mendelian randomization (MR)^64^ framework^30,65^ to test for causal effects of i) nine protein mediator candidates identified in the mediation framework and the corresponding 26 significant brain MRI traits and ii) IDP-associated proteins on cardiovascular (arrhythmias, CAD, HF, hypertension, stroke) and neurological/psychiatric outcomes (dementia, neurodegenerative diseases, psychiatric disorders). Instrumental variables were *cis*-pQTLs (within ±1 Mb of the gene) from the UKB-PPP^20^ at genome-wide significance (*p*<5×10⁻⁸). Variants were clumped for linkage disequilibrium (LD) (r²<0.01) using PLINK (version 1.9)^66^ and a 1000 Genomes European reference^67^ panel, discarding instruments with F-statistic<10 to mitigate weak instrument bias.

75 GWAS summary statistics for eight disease categories and 26 significant available brain MRI traits GWAS were drawn from the OpenGWAS project^29^, integrating data from substantial cohorts including the NHGRI-EBI Catalog^46^, MRC IEU OpenGWAS^29,30^, and FinnGen^47^, typically involving >10,000 individuals of European ancestry. pQTLs and outcome GWAS data were harmonized to align effect alleles, using proxy SNPs (LD r^2^>0.8) when necessary. Twelve MR methods, including inverse-variance weighted (IVW) method (fixed effect, multiplicative random effects with/without corrected standard error under dispersion), MR-Egger regression, Simple median, Weighted median, Penalised weighted median, Simple mode, Simple mode (NOME), Weighted mode, Weighted mode (NOME), were applied. For single-variant instruments, the Wald ratio was used. Heterogeneity was assessed via Cochran’s Q test and the I² statistic, with MR-Egger to test for pleiotropy. We used 5% FDR corrected *p*-values on IVW to define robust causal inferences. Additionally, Steiger filtering was employed to verify the directionality of associations between proteins and outcomes, with *p* < 0.05 indicating that the effect direction is from exposure to an outcome. Proteins on sex chromosomes or lacking significant *cis*-pQTLs were excluded. For the disease MR analysis, this main analysis comprised 327 CMR IDP-specific, 36 BMR IDP-specific associated proteins, and 35 proteins that associated with both CMR IDP and BMR IDP.

To validate and replicate our findings in disease MR analysis, we repeated the MR analyses using plasma proteomics data from the deCODE^52^ and Fenland^53^ studies, both measured using the SomaScan platform. The deCODE dataset included 4,907 aptamers measured in 35,559 Icelandic individuals, and the Fenland dataset included 4,775 aptamers measured in 10,708 European participants. The same MR pipeline was applied to assess causal associations between proteins and disease outcomes. Overlaps in significant protein-outcome associations between the UKB proteomics dataset and the deCODE/Fenland datasets were quantified to evaluate the consistency of findings across cohorts.

### Genetic colocalization analysis

A Bayesian colocalization approach^68^ was used to detect shared causal variants between protein *cis*-pQTLs (UKB-PPP^20^) and GWAS hits for eight cardiovascular, neurological, and psychiatric conditions matching the MR targets. We retrieved GWAS data from OpenGWAS^29,30^ for studies with sample sizes >10,000 and with significant findings (*p*<5×10⁻⁸).

Significant loci were defined by pQTL signals at *p*<5.0×10⁻⁸, LD-clumped at r^2^<0.1 within ±1 Mb of the lead variant. Loci with ≥50 overlapping SNPs in the pQTL and GWAS summary statistics and ≥1 genome-wide significant SNP (*p*<5.0×10⁻⁸) for both pQTL and GWAS were tested via the coloc R package^68^ with default priors (*p*_1_ = 1 × 10^−4^, *p*_2_ = 1 × 10^−4^, *p*_12_ = 1 × 10^−5^). A secondary analysis used a more stringent *p*_12_ = 5 × 10^−6^. There are four Posterior probabilities hypothesis (PP.H), PP.H0, a genetic variant in the region is not associated with either trait; PP.H1, a genetic variant in the region is associated with Trait 1 (pQTL) but not Trait 2 (disease GWAS); PP.H2, a genetic variant in the region is associated with Trait 2 (disease GWAS) but not Trait 1 (pQTL); PP.H3, a genetic variant in the region is associated with both Trait 1 (pQTL) and Trait 2 (disease GWAS) but with different causal variants; PP.H4, a genetic variant in the region is associated with both Trait 1 (pQTL) and Trait 2 (disease GWAS) and with the same causal variant. We considered PP.H4 > 0.8 as high support and PP.H4 > 0.5 as moderate support for colocalization, indicating that the pQTL and disease GWAS signals are likely driven by the same underlying causal variant rather than by linkage disequilibrium.

### Biomarker and drug-target reference

We cross-referenced candidate proteins with MarkerDB (December 2023 release)^45^ to identify diagnostic protein biomarkers. MarkerDB collates diagnostic, prognostic, exposure, and predictive biomarkers across 670 human conditions. For drug targets, we consulted DrugBank (March 2024 release, version 5.1.12)^48^, noting any approved therapeutics that act via these proteins. Proteins serving as bona fide therapeutic targets or recognized off-target interactions were flagged accordingly.

## Data availability

All UK Biobank data (demographics, imaging, proteomics) are accessible via application to the UK Biobank. Raw MRI data and derived IDPs are also released to approved researchers. pQTL summary statistics can be assessed from the UKB-PPP study (https://metabolomips.org/ukbbpgwas/), the deCODE Health study (https://www.decode.com/summarydata/), and the Fenland study (https://www.omicscience.org/apps/pgwas). Single-cell RNA-seq data are available on GEO (GSE155468, GSE253903).

## Code availability

All analyses and visualizations were implemented in R (v4.4.0) and Python (v3.12.0). Scripts are available upon publication, including those used to replicate the analyses and figures presented in this manuscript.

## Acknowledgements

This study utilized data from the UK Biobank resource under application numbers 197947 and 88365. We sincerely thank the participants and staff of the UK Biobank for their invaluable contributions. We also gratefully acknowledge the participants and investigators of the FinnGen study. This work was supported by grants from the U.S. National Institutes of Health (NIH) (R01 AG061034 and R35 HL155318 to A.R., K08 HL177169 to S.A.K.), the Burroughs Wellcome Fund (Fund ID 1416136 to S.A.K.), and the American Heart Association (AHA) (23MERIT1038415 and 24SFRNPCN1284382 [URLs: https://doi.org/10.58275/AHA.24SFRNPCN1284382.pc.gr.194135 and https://doi.org/10.58275/AHA.24SFRNCCN1276092.pc.gr.194131] to A.R.).

## Author contributions

A.R. conceived the study, supervised the research, and revised the manuscript. C.W. designed the study, collected the data, performed the analyses, and drafted the manuscript. D.T.L. contributed to data analysis, project discussions, and manuscript revisions. M.W. provided feedback on the study design and statistical methodology and revised the manuscript. H.Y.L. provided access to datasets and offered guidance on statistical methods. C.Y.L. performed ligand-receptor interaction mapping. Z.X.Y. and J.Q.H. performed the single-cell RNA sequencing and cell-type–specific analyses. J.R.B.G. supported pathway and biological process analyses. S.Y.H. developed the interactive web portal for data visualization. Q.L.Z. and M.X.Q. assisted with manuscript editing and figure preparation. S.A.K. performed the animal experiments and revised the manuscript.

## Additional Information

**Extended data** is available for this paper.

---

**Extended Data Table 1.**
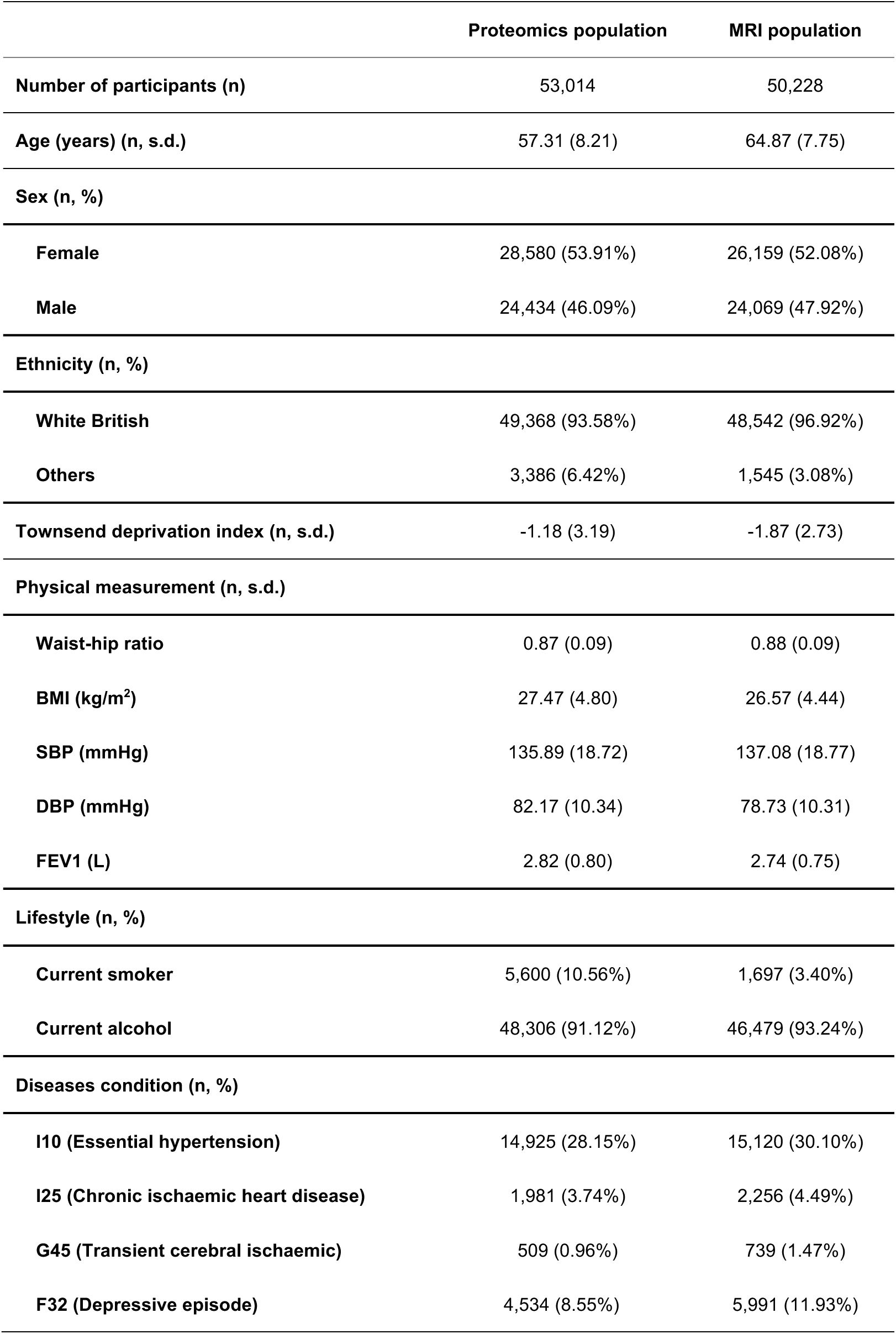
Main demographics of the participants.

**Extended Data Fig. 1.**
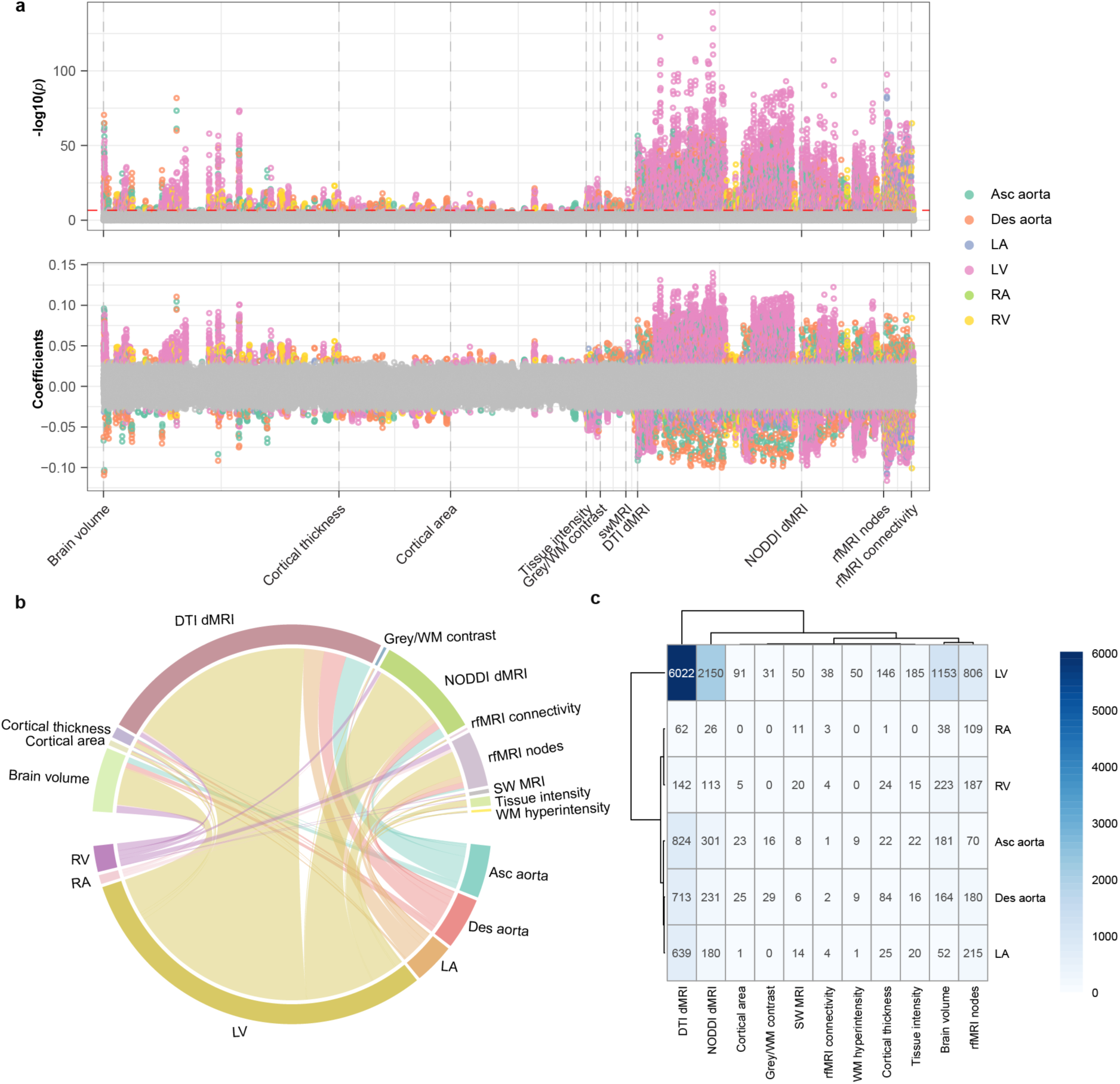
Univariate correlation between CMR and BMR traits. **a.** Manhattan plot of Pearson correlations between 82 CMR and 2,222 BMR traits. The x-axis shows BMR trait categories; the y-axis shows −log_10_(*p*) and correlation coefficients. The red line denotes the Bonferroni-corrected threshold (*p*<2.74×10^−7^). Gray points represent negative correlations. CMR traits are color-coded. **b**. Significant correlations (Bonferroni-corrected) between CMR and BMR trait categories. **c**. Counts of significant CMR-BMR correlations. WM, white matter; swMRI, susceptibility-weighted MRI; DTI, diffusion tensor imaging; NODDI, neurite orientation dispersion and density imaging; dMRI, diffusion-weighted MRI; rfMRI, resting-state functional MRI; LV, left ventricle; LA, left atrium; RV, right ventricle; RA, right atrium; Asc aorta, ascending aorta; Des aorta, descending aorta; CMR, cardiac MRI; BMR, brain MRI.

**Extended Data Fig. 2.**
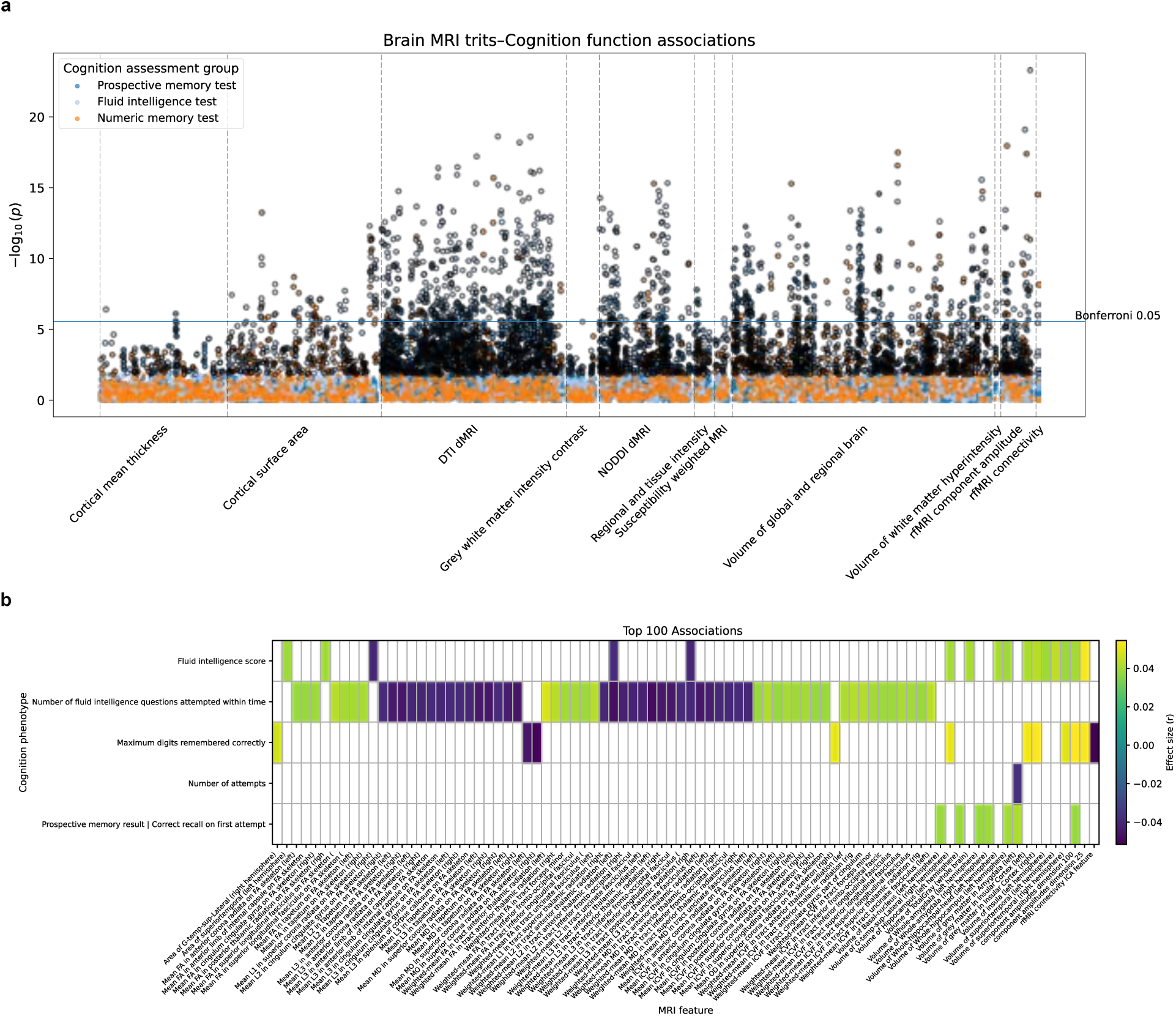
Univariate associations between cognitive assessments and brain MRI IDPs. **a.** Manhattan plot of Pearson correlation results between three cognitive assessment domains and 2,222 brain MRI imaging-derived phenotypes (BMR IDPs). The x-axis indicates BMR IDP categories and the y-axis shows −log_10_(*p*). The horizontal blue line marks the Bonferroni-corrected significance threshold (*p*<2.81×10⁻⁶). Points outlined in black denote correlations that remain significant after FDR correction. **b.** Heatmap of the top 100 most significant (Bonferroni-corrected) cognition-BMR IDP associations. Colors represent the Pearson correlation coefficients.

**Extended Data Fig. 3.**
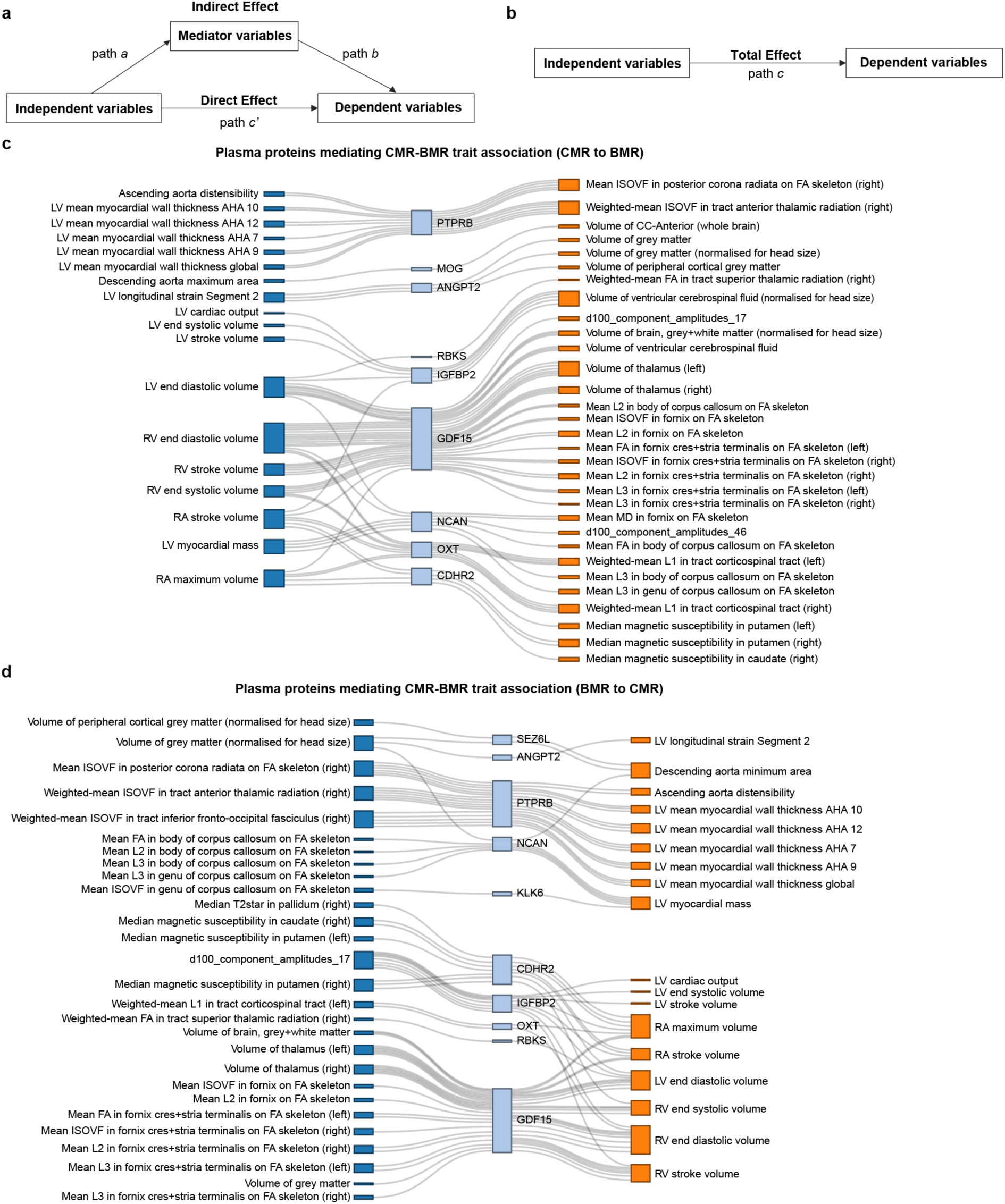
Mediation analysis of proteins on CMR-BMR trait interactions. **a–b.** Diagram of mediation models with (**a**) and without (**b**) the mediator. The total effect (path c) is the impact of the independent MRI trait on the dependent MRI trait; the direct effect (c′) remains after adjusting for the mediator; and the indirect effect (path *a* × path *b*) is mediated through the protein. **c–d**. Plasma proteins that significantly mediate interactions from CMR to BMR traits (**c**) or from BMR to CMR traits (**d**). ISOVF, isotropic volume fraction; FA, fractional anisotropy; MD, mean diffusivity.

**Extended Data Fig. 4.**
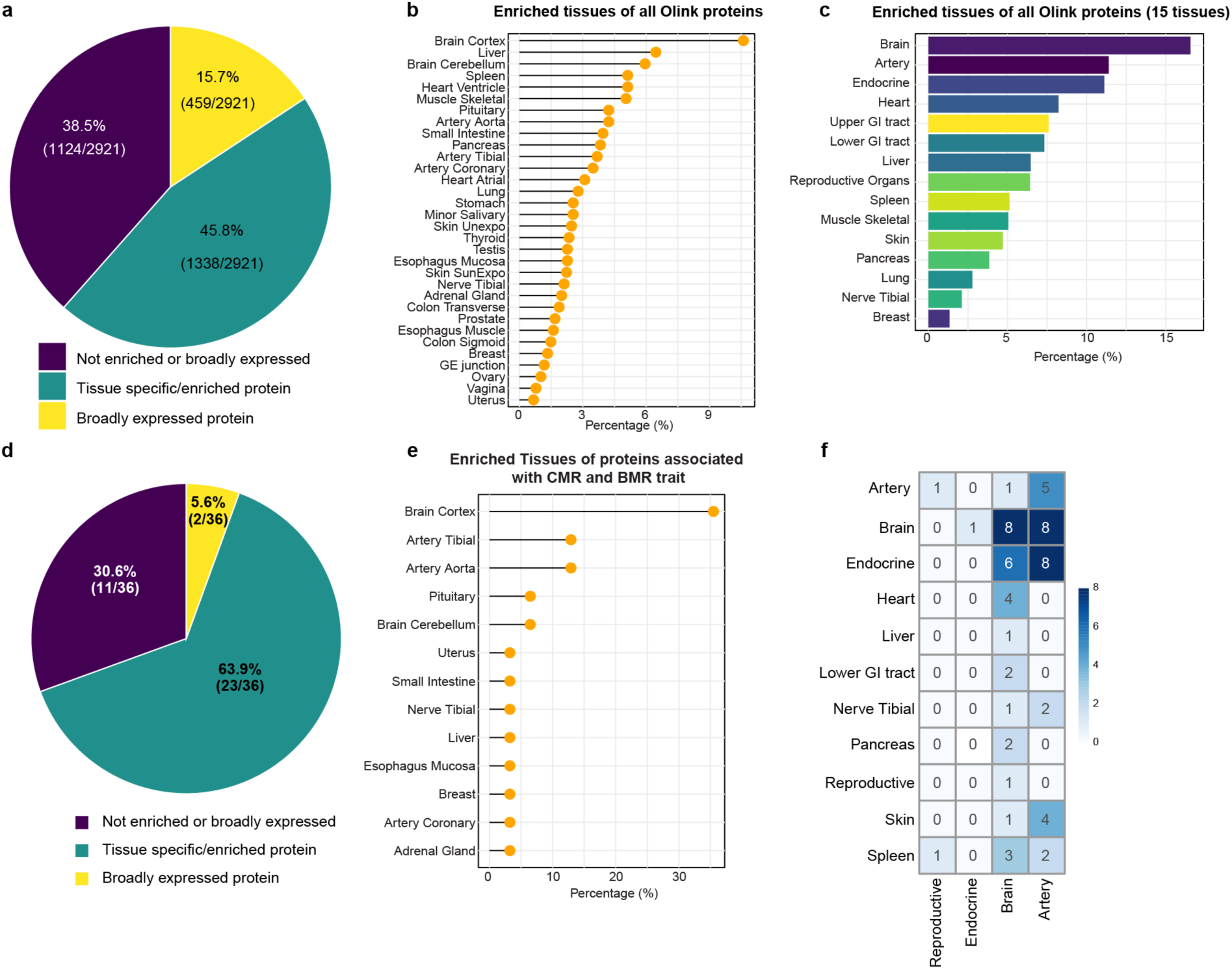
Tissue enrichment of all Olink proteins and proteins associated with both CMR and BMR trait. **a.** Distribution of tissue-enrichment types for all 2,922 Olink proteins. **b–c**. Tissue enrichment of tissue-enriched and tissue-specific proteins across 32 GTEx tissues (**b**) and consolidated into 15 tissue groups (**c**). **d–f.** Tissue enrichment for proteins associated with both CMR and BMR traits. **d**. Distribution of tissue-enrichment types. **e**. Tissue enrichment across 32 GTEx tissues. **f.** Quantitative ligand-receptor connections; the x-axis represents ligands, and the y-axis represents receptors.

**Extended Data Fig. 5.**
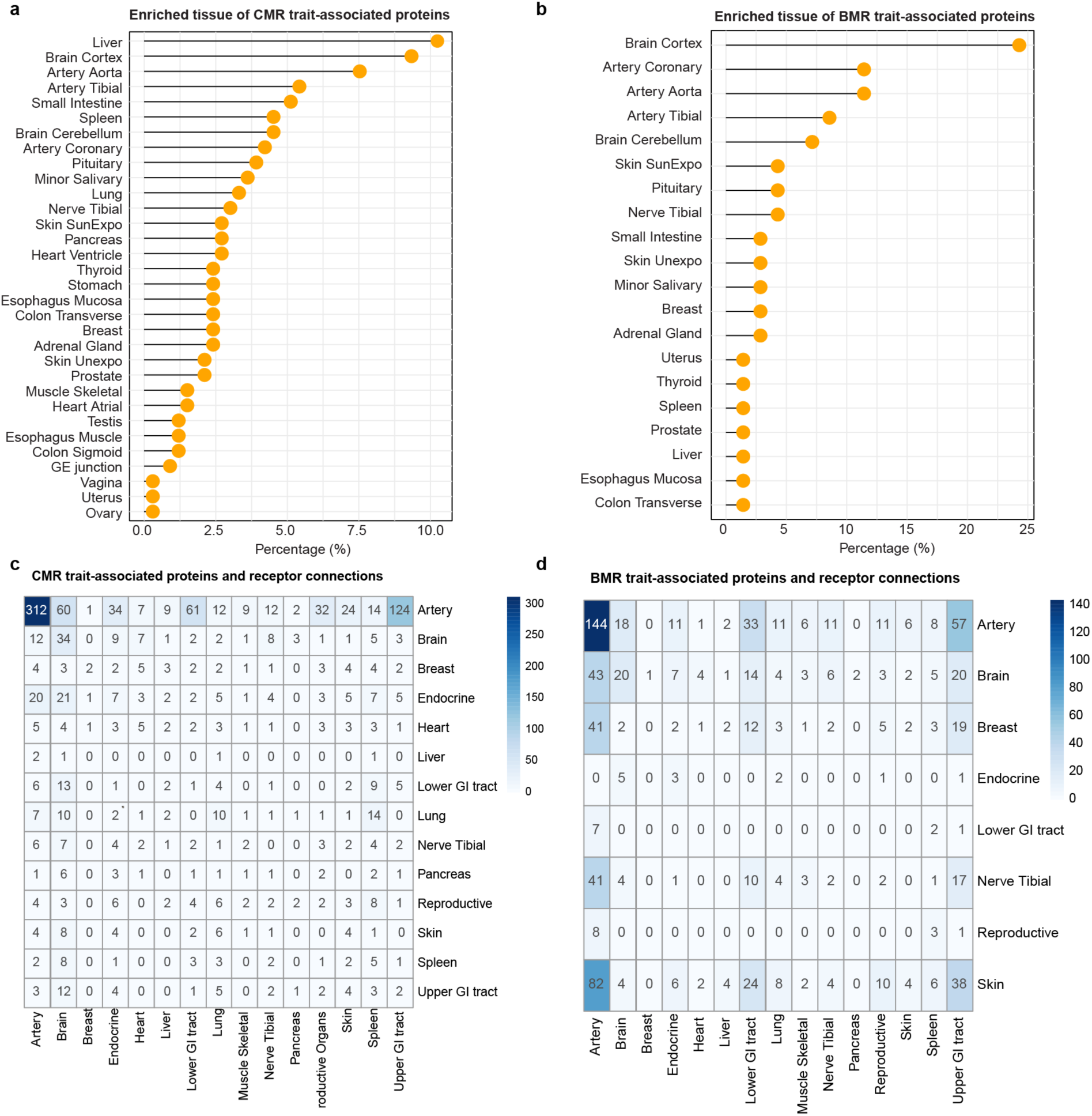
MRI-associated circulating protein tissue enrichment. **a–b.** Tissue enrichment for tissue-enriched and tissue-specific proteins associated with CMR traits (**a**) and BMR traits (**b**) across 32 GTEx tissues. **c–d**. Quantitative ligand-receptor mapping for CMR (**c**) and BMR (**d**) trait-associated proteins. The x-axis shows receptors, and the y-axis shows ligands.

**Extended Data Fig. 6.**
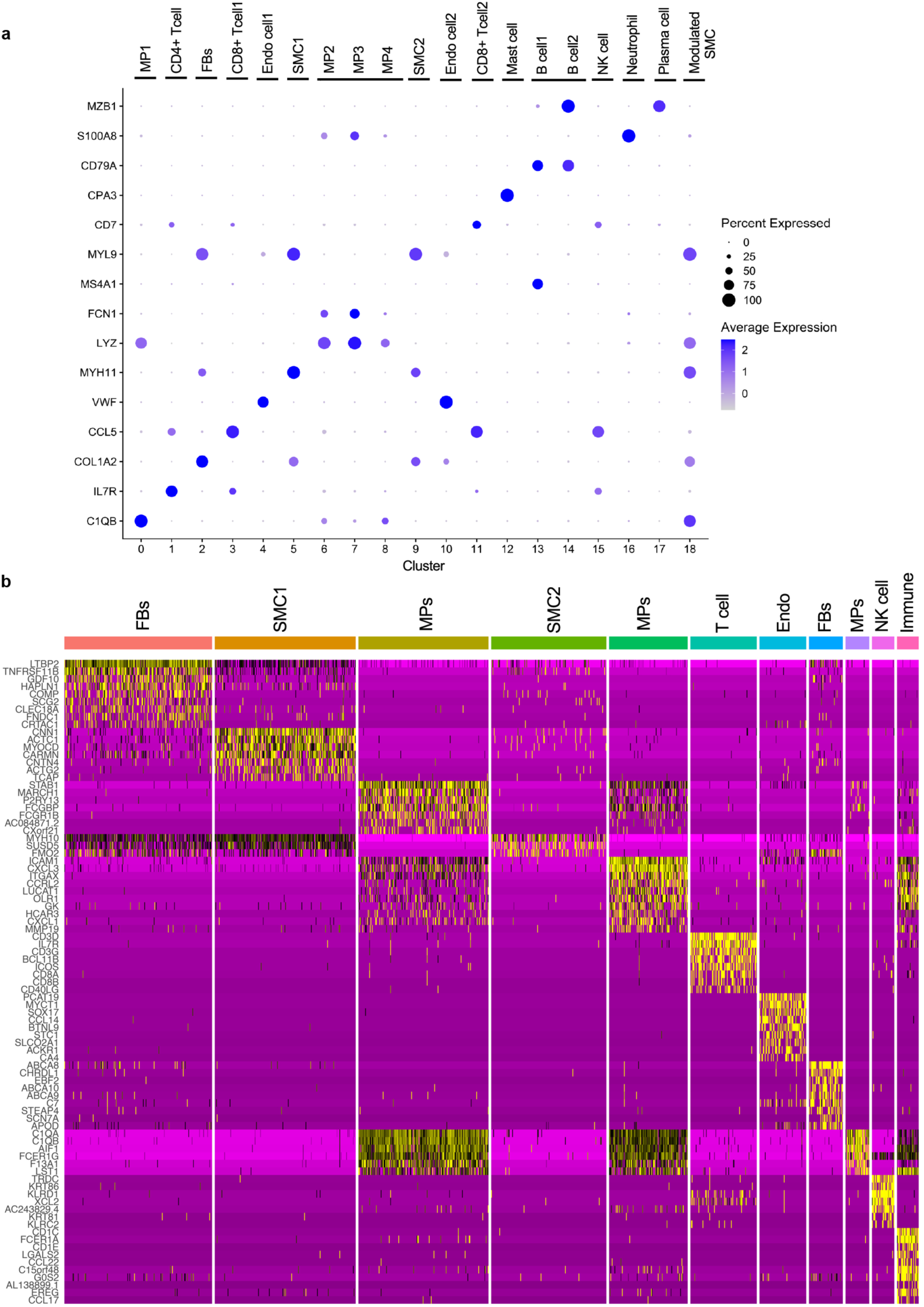
Cell-type markers in artery tissue. **a–b.** Key cell-type biomarkers identified in single-cell RNA-sequencing data from carotid (**a**) and aorta (**b**) tissues, highlighting markers for fibroblasts (FBS), smooth muscle cells (SMCs), endothelial cells, and immune cells.

**Extended Data Fig. 7.**
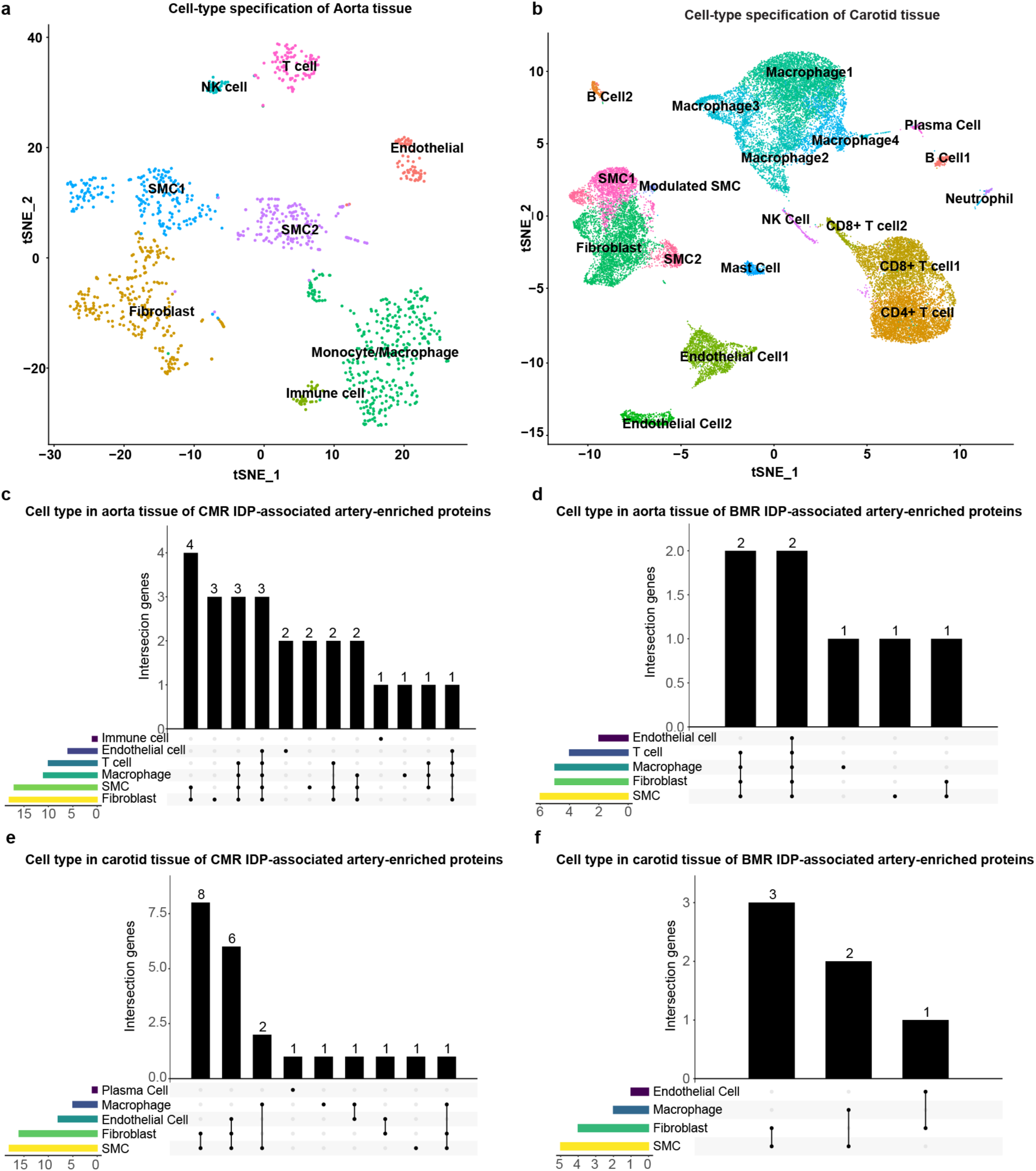
Cell-type specificity of candidate proteins in artery tissue. **a–b.** Single-cell clustering of aorta (**a**) and carotid (**b**) tissues. **c–d**. Counts of cell-specific genes corresponding to artery-enriched, CMR trait-associated proteins (**c**) or BMR trait-associated proteins (**d**) in the aorta. **e–f**. Counts of cell-specific genes corresponding to artery-enriched, CMR trait-associated proteins (**e**) or BMR trait-associated proteins (**f**) in the carotid. SMC, smooth muscle cell.

**Extended Data Fig. 8.**
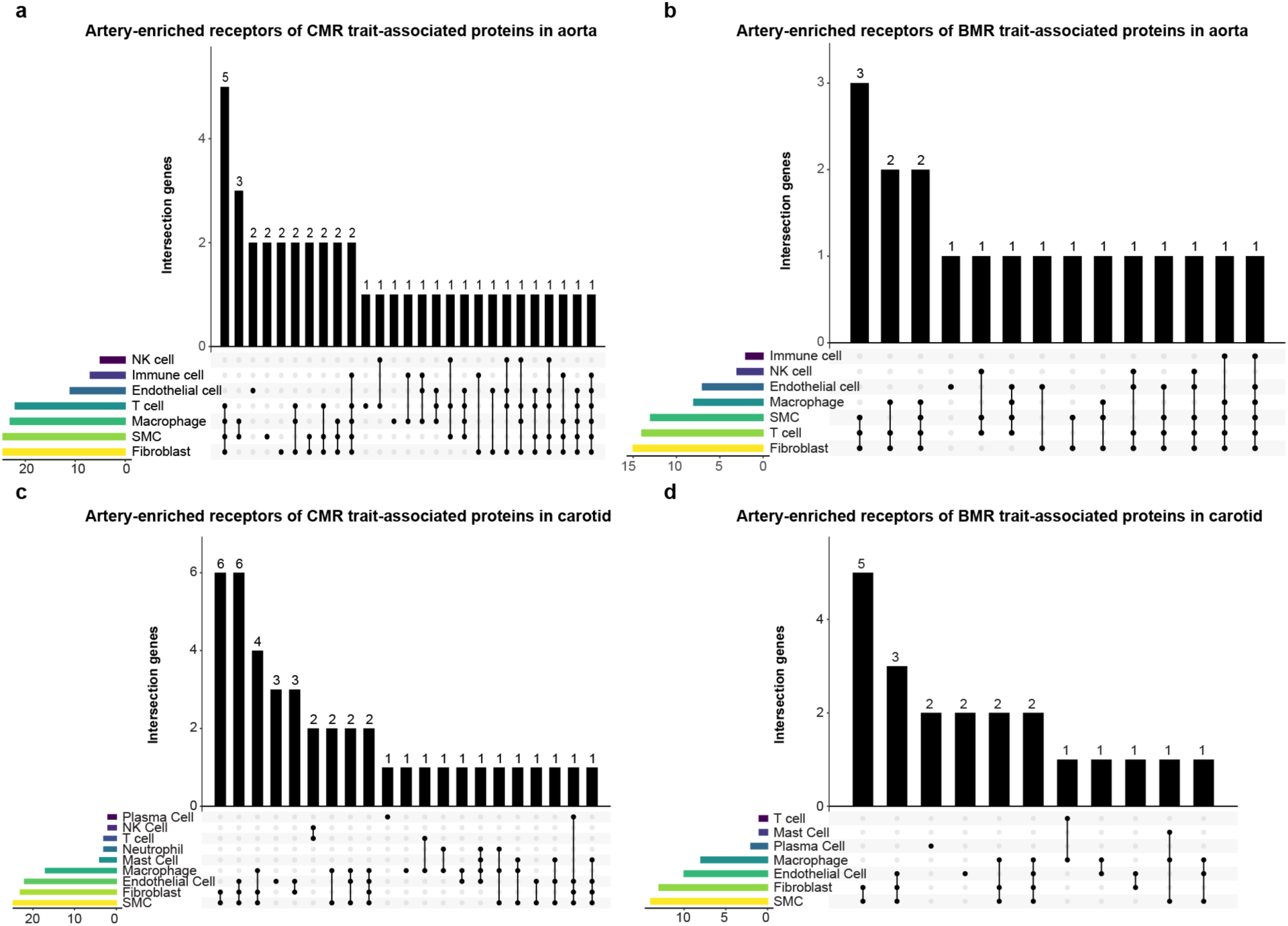
Cell-type specificity of artery-enriched receptors. **a–b.** Counts of cell-specific genes corresponding to artery-enriched receptors for CMR trait-associated (**a**) or BMR trait-associated (**b**) ligands in the aorta. **c–d**. Counts of cell-specific genes corresponding to artery-enriched receptors for CMR trait-associated (**c**) or BMR trait-associated (**d**) ligands in the carotid.

**Extended Data Fig. 9.**
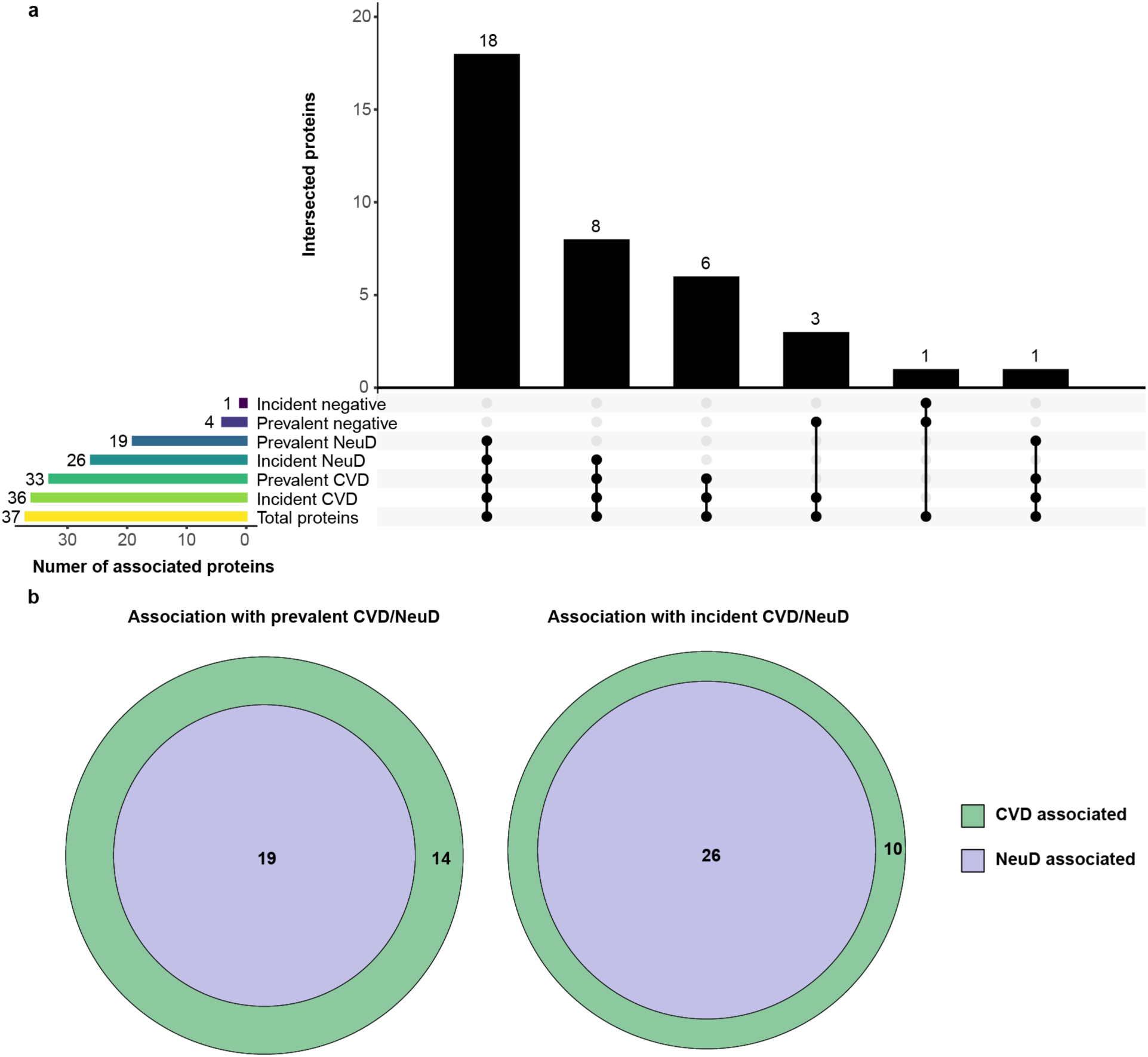
Associations between prevalent and incident disease and proteins associated with both CMR and BMR trait. **a.** Proteins associated with incident and prevalent CVD and NeuD among proteins associated with both CMR and BMR trait. **b**. Overlap of proteins associated with incident/prevalent CVD and NeuD among proteins associated with both CMR and BMR trait. CVD, cardiovascular diseases; NeuD, neurological diseases.

